# Genetic Mutations Associated with Hormone-Positive Breast Cancer in Ethiopian Women

**DOI:** 10.1101/2020.11.25.20238881

**Authors:** Alyssa D. Schwartz, Afua Adusei, Solomon Tsegaye, Christopher A. Moskaluk, Sallie S. Schneider, Manu O. Platt, Daniel Seifu, Shelly R. Peyton, Courtney C. Babbitt

## Abstract

In Ethiopia, a breast cancer diagnosis is associated with a prognosis significantly worse than that of Europe and the US. Further, patients presenting with breast cancer in Ethiopia are far younger, on average, and patients are typically diagnosed at very late stages, relative to breast cancer patients of European descent. Emerging data suggest that a large proportion of Ethiopian patients have hormone-positive (ER ^+^) breast cancer. This is surprising given 1) the aggressive nature of the disease, 2) that African Americans with breast cancer frequently have triple negative breast cancer (TNBC), and 3) these patients typically receive chemotherapy, not hormone-targeting drugs. To further examine the similarity of Ethiopian breast tumors to those of African Americans or of those of European descent, we sequenced matched normal and tumor tissue from Ethiopian patients from a small pilot collection. We identified mutations in 615 genes across all three patients, unique to the tumor tissue. Across this analysis, we found far more mutations shared between Ethiopian patient tissue and White patients (103) than we did comparing to African Americans (3). Several mutations were found in extracellular matrix encoding genes with known roles in tumor cell growth and metastasis. We suggest future mechanistic studies on this disease focus on these genes first, toward finding new treatment strategies for breast cancer patients in Ethiopia.

## Introduction

Women in Ethiopia develop extremely aggressive breast cancer, often at a very young age (1). The most comprehensive study followed 1,070 women and determined that two key contributors to an increased hazard ratio were 1) being diagnosed younger than 50 years old, and 2) being diagnosed with stage 3 breast cancer. In the US, women are determined to be high risk patients if they have family history of breast cancer (3, 4), high breast density (5), or a mutation in a known high-risk gene, such as BRCA1/2 (6). While the lack of information pertaining to family history and breast density (access to mammograms) is a public health issue faced by many populations around the globe, only 30% of familial breast cancers can be traced to a known genetic mutation (7). That number is even lower in non-western populations: *e*.*g*. a Korean study found less than 22% of familial breast cancer was traced to a BRCA1/2 mutation (8).

Breast cancer is the most common form of cancer diagnosed in Ethiopian women (31.5 - 33% of cancers in women) (9, 10). Kantelhardt *et al*. analyzed 352 patients treated at Addis Ababa University hospital and found that only 31% of tumors were hormone receptor negative. This is in stark contrast to related publications on patients from West Africa, which were up to 76% hormone receptor negative (11). This is an important point, as studies in the US have largely focused on the fact that aggressive African American disease is largely triple negative (TNBC). The overarching challenge here is the limited information on this hormone-positive East African disease (1). This is critically important, as Ethiopian women with breast cancer from rural areas have a 50% survival rate after 2 years (12). There are many possible reasons why Ethiopian women present with aggressive, hormone-positive breast cancer at an early age, such as environmental factors, age of menarche, socioeconomic status, healthcare resources, stigma, and distance to major hospitals (13, 14).

There is a paucity of studies on the genetics of breast tumors of African women in general, and even less of those of East Africans. Most studies have focused on documenting mutations of the BRCA1/2 genes (15). This is particularly important when trying to relate this disease to that of TNBC in African Americans. However, we are not aware of any studies that have performed whole exome sequencing on patients within an East African or Ethiopian community. This is the critical data that we need to find the driver genes of breast cancer in Ethiopian women, and to determine if these driver genes are unique to this population or not.

## Results

### Ethiopian women present with aggressive ER1 breast cancer at a young age

Through discussions with surgeons and oncologists at the Black Lion Hospital in Addis Ababa, we were alerted to the fact that Ethiopian women are diagnosed at a much younger age than women of European descent. A recent study found that the overall mean age at diagnosis was 43 years old, and 40% of patients were under 40 years old (16). Amongst this cohort, patients with Luminal B breast cancer had the youngest median age at diagnosis at 35, followed by Her2-enriched at 41, Triple negative at 46, and Luminal A at 47. The age of diagnosis across breast cancer subtypes is consistently younger for those born in Africa compared to those born in the US (Figure 1) (2, 17). Also, at the time of diagnosis, Ethiopian patients had a significantly higher chance of having a higher-grade pathology compared to White and African Americans, across all subtypes (17). Second, a recent study from Jemal and Fedewa demonstrated that breast cancer cases in East Africa (74% of these are from Ethiopia) are more likely to be ER^+^ compared to West African or African American cases (Figure 1) (2).

**Figure 1:**
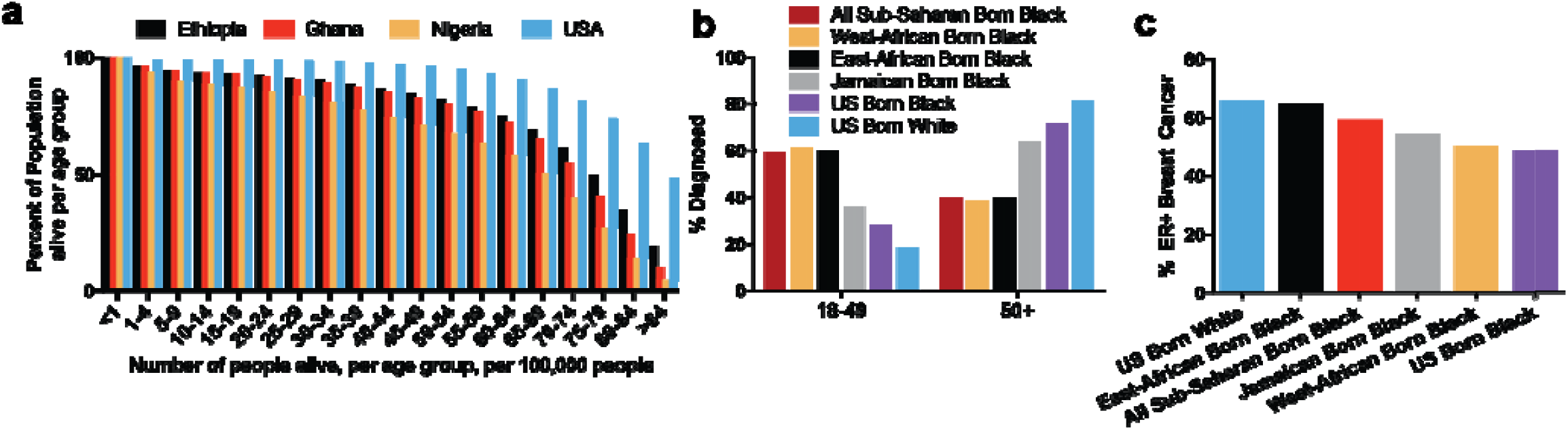
Age and subtyping data for Ethiopian women with breast cancer. a) Data from world health organization on mortality data across populations from Ethiopia (black), Ghana (red), Nigeria (yellow) and US (blue). b) Data adapted from Jemal and Fedewa on age of diagnosis of breast cancer across relevant subpopulations (2). c) Proportion of ER+ breast cancer patients from the same reference.

### Whole exome sequencing reveals unique mutations in Ethiopian breast tumors

We performed DNA sequencing on three tumors and healthy tissue from matched patients. Each of the three tumors were identified as ER^+^ using immunohistochemical subtyping. We identified mutations in 615 genes across all three patients, unique to the tumor tissue (Table S1). Two patients had a mutation in a different spot in the BRCA2 gene, which is not surprising based on the number of variants of uncertain significance in BRCA2 (18).

Additionally, data from this population shows that patients with a family history were more likely to be ER-than patients without a family history (19), suggesting that many of these patients’ cancers are due to somatic mutations. We also searched for mutations in genes related to breast cancer aggression, subtype, and progression. Gene ontology enrichments (20) showed MSigDB Oncogenic categories (21) related to KRAS, PTEN, and EGFR signaling (Table 1), demonstrating that at least some of the somatic variants in population are in the same pathways as other ER ^+^ tumors in other populations (22, 23).

**Table 1.**
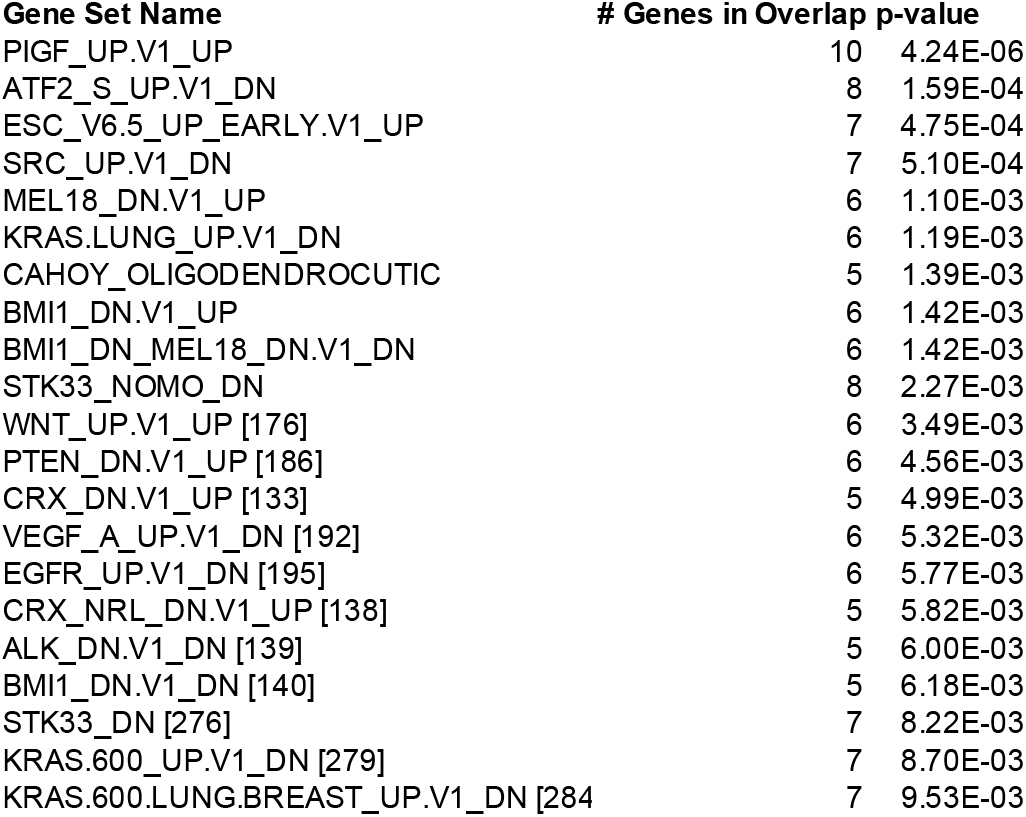
GSEA enrichments of the SNPs found across the Ethiopian patient tumors.

### Overlap between mutated genes in Ethiopian samples and the TCGA

In our preliminary analyses of our Ethiopian whole exome sequencing (WXS) and data from the TCGA we see a larger overlap (Figure 2) in mutation in the genes also identified as mutated in Caucasian samples, as compared to the overlap with TCGA African American mutations (103 vs. 3 genes, Figure 2, Table S2, S3). These are mutations in genes that are not seen in our normal tissue samples. Our list of genes with mutations was also much smaller compared to the extensive data from the TCGA (276 as compared to 16,897 in the White patients, and 9,073 in African Americans). Preliminary analysis from a small number of Ethiopian tumors suggests that, on a genetic level, the mutational spectrum of tumors from ER1 Ethiopian tumors are more similar to that of White women in the US than to African Americans.

**Figure 2.**
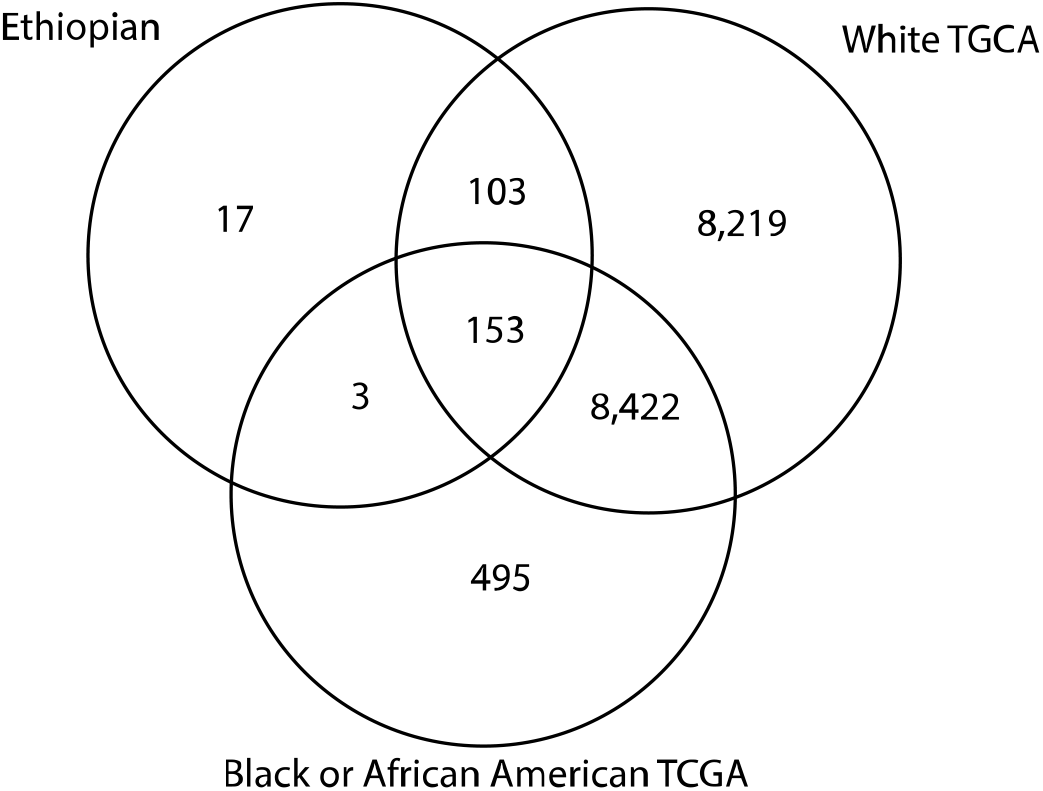
Overlaps in genes showing mutations in breast cancer patients from the Ethiopian population and White or African American data from the TCGA.

Gene ontology enrichment analysis of the 103 overlapping shows two ontogenetic signature categories “Genes down-regulated by treatment with VEGFA” and “Genes up-regulated by treatment with PIGF”. GO ontology enrichments show that these are enriched for categories in “Cytoskeletal protein binding”, “Ras protein signal transduction”, and “Plasma membrane protein complex” (Table S2).

Given that these were ER+ tumor samples, it was not surprising that we uncovered several genes implicated in ER signaling. We identified mutations in tumor suppressor gene, NBL1, which has been shown to be upregulated in response to an estrogen mimic (24). Additionally, we found mutations in multiple members of the protein tyrosine phosphatase (PTP) family of kinases, of which several members have been implicated in hormone therapy resistance. An additional gene of note is the oncogene VAV3, which is activated downstream of ER signaling via PI3K phosphorylation (25). Together these suggest that the higher incidence of ER1 tumors in Ethiopian women may be linked to hormone-specific mechanisms of transformation and progression.

### We found mutations in ECM and ECM-related proteins in all tumor samples

We found 615 genes that had mutations across all three patients (but were not mutated in the paired healthy tissues for any of the women, Table S1). We were particularly interested in 5 of these genes that code for proteins in the extracellular matrix (ECM): LAMA5, LAMC3, COL6A5, MUC12, or ECM-downstream signaling proteins: MAP2K3, because of the known role of the ECM in tumor growth and metastasis (26-30). We speculated that mutations in these genes could be partially responsible for the documented aggressive nature of breast cancer in Ethiopia.

LAMA5 is the gene that encodes for the alpha-5 subunit of laminin-10 (laminin-511), laminin-11 (laminin-521), and laminin-15 (laminin-523). LAMA5 is normally highly expressed in the skin, placenta, and lung, and it has been studied primarily for its role in stem cell maintenance. The few LAMA5 papers possibly related to our observation include reports on how the secreted protein can support breast cancer cell survival (31), adhesion of epithelial cells (32), melanoma cell motility (33), and chemotaxis of macrophages (34). LAMC3 is laminin subunit 3, which is highly expressed in the placenta and testes, and it was recently identified through another GWAS study as linked to oral and pharyngeal cancer (35). COL6A5 is a member of the collagen superfamily, and it is highly expressed in the skin and lung. Mutations in this gene are significantly associated with itch, eczema, dermatitis, etc. Although not directly related to breast cancer, clearly it is involved in epithelial cell detachment, and so its mutation here could be related to primary tumor cell detachment and ultimately metastasis.

MUC12 encodes an integral membrane glycoprotein (mucin). Mucins are O-glycosylated proteins that form protective mucous barriers on epithelial surfaces. MUC12 is specifically expressed in the colon, and it is linked to colitis and aggressive colon cancer, so its appearance in our study is both novel and puzzling. Future studies with a larger patient cohort are critical to determining the link between mutations in the intestine, colon, and breast. Finally, MAP2K3 (mitogen-activated protein kinase kinase 3) is activated during stress and is notably involved in oncogene activation. It is expressed in all tissues, most highly in the bone marrow. Finding mutations in this gene was not surprising, given its known role in breast cancer (36-39).

## Discussion

The vast majority of breast cancer patients in Ethiopia are treated with radiotherapy and broad-spectrum chemotherapy, resulting in many toxicity-related deaths. Additional immunohistochemical (IHC) and genetic analyses could reveal improved treatment strategies for these patients. For example, if a large proportion of these patients have ER1 (luminal) breast cancer, then tamoxifen would be an obvious course of front-line treatment. This question was first explored by Jemal and Fedawa, finding that Ethiopian women had much higher ER+ breast cancer compared to Western Africa born women (2). This likely spurred AstraZeneca to start such a tamoxifen trial (1), but there is no long-term plan in place. One of the few publications on Ethiopian breast cancer found that 64% of the patients in their analyzed cohort were indeed hormone receptor-positive (11). Tamoxifen is a generic drug that revolutionized treatment in the US in the 1970s (40), and adding this to the treatment portfolio in Ethiopia could radically transform the disease in this country.

Ethiopia has the 13^th^ highest population in the world, and up to 85% of the population lives in rural areas (41). There has been a recent increase in the use of chemical pesticides at farms in Ethiopia (42), and many of these pesticides have been identified as endocrine disrupting chemicals (similar to BPA which is used in plastic production and epoxy resins), which has been mirrored by an increase in endocrine-related diseases, including, but not limited to, breast cancer (43, 44) and thyroid disorders (45, 46). The specific mechanisms of how environmental factors put Ethiopian women at risk for breast cancer is outside the scope of this study, but the mutational information we identified here could provide clues for future investigation.

Genetic information, such as that from DNA sequencing could detect genetic risk factors, and high penetrance mutations in understudied populations could identify new therapeutic targets. This has previously been done with an activating mutation in ER^+^ breast cancer (47), but focused on populations primarily of European descent. Genetic analysis of a small population with non-BRCA familial history of breast and ovarian cancer identified mutations in several genes involved in DNA repair, cell signaling, and apoptosis that were possible predictors for heritable cancer risk (48, 49).

In the US, whole exome sequencing has been used to identify single nucleotide polymorphisms (SNPs) in exome regions associated with risk for cancers in a patient-specific manner (49). Identification of prevalent mutations will lay the groundwork for identifying risk factors or targeted drugs. Additionally, we and others can use this mutation database to develop new exome sequencing panels for breast cancer in women of East African descent. One interesting finding we had here is the ontogenetic signature for genes up-regulated in response to PIGF (Phosphatidylinositol-glycan biosynthesis class F protein). PIGF is more often upregulated in triple negative breast cancer, so it is striking that we observed an increase in this pathway in these ER1 breast cancers. PIGF has been noted as a biomarker in South African pancreatic ductal adenocarcinoma (50). PIGF signaling affects both stemness of cancer cells (51), as well as macrophage polarization (52), which could increase the aggressiveness of the disease.

It is surprising to us that all three of these patients harbored somatic mutations in multiple “risk factor genes” denoted by these panels. At the same time, all the mutations we identified were 1) *specific to* the tumor tissue, and 2) *not found* in the matched healthy tissue we analyzed. This points toward these mutations, at least for this small initial patient cohort, to not be reliable risk factor mutations. We focused here on ECM and ECM-related proteins because of the known role of the ECM environment in driving breast cancer growth, invasion, and metastasis (28, 29, 53-59). Unfortunately, we cannot test directly whether mutations in these proteins are directly responsible for the aggressive nature of the disease in these women. Regardless, the overlap with these shared mutations in the Ethiopian samples and those from ER+ samples in White women from the TCGA data set is striking. It is interesting to note that none of the mutations we saw in all three of our tumor samples were highlighted in a recent review of mutations characteristic of tumors in African women (15). This perhaps points to a mutational spectrum unique to tumors in Ethiopia, but we can only speculate as to whether these mutations are relevant to the aggressive phenotypes observed.

One study that stands out is from the Howard University Hospital and examines confirmed breast cancer cases in African American women. Of note, the Washington D.C. area has a high proportion of women of Ethiopian descent. This group found that having an induced abortion, which allowed for part of the normal estrogen cycle during pregnancy (60) was a significant risk factor for developing breast cancer. This correlation was not present for women who miscarried, which is associated with lower estrogen levels (61). This does not appear to be a universal finding, so the higher fraction of Ethiopian women likely represented in this work may be important.

Breast cancer in African American women is notorious for having an overly aggressive phenotype, and it disproportionately affects younger women (62). Within the subgroup of these patients that have inherited disease, many of them have mutations in BRCA1, BRCA2, PTEN, and TP53. A recent study of about 300 African American patients with invasive disease showed that about a third of these patients were of the triple negative breast cancer (TNBC) subtype, with another third ER^+^. However, the limited data indicate that this population is predominately of West African descent. Yet, multiple lines of evidence lead one to the hypothesis that Ethiopian breast cancer is unlike the African American disease. First, a substantial amount of patients at the Black Lion hospital are presenting with cancer at a far younger age than typical for African American disease (nearly 80% of patients are under 50 (1)), and women are disproportionately presenting with advanced disease (greater than 70% are stage 3-4, (1)). Second, a recent study from Jemal and Fedewa demonstrated that breast cancer cases in East Africa (74% of these are from Ethiopia) are more likely to be ER^+^ compared to West African or African American cases (Figure 1) (2). Third, aggressive ER^+^ breast cancer in the US is relatively rare, even for African American patients (63), so it is highly likely that the driving factors, genetic or otherwise, for this disease in Ethiopia is distinct from that of African Americans. Outside of our study, it would be valuable to compare the genomic data we have here to White women with ER1 breast cancer in the US, to White women in the US with TNBC, and to African American women with TNBC. Unfortunately, that is not yet possible to do with the publicly available data in the TCGA, and the data we present in Figure 2 is across all subtypes.

Genetic mutations that contribute to the development of cancer are characterized by their penetrance, which is the proportion of individuals carrying a mutation that also show the associated phenotype. Current studies suggest that any non-BRCA familial mutations are not high-penetrance, which would explain why no other variants have been found and explored as extensively (64). The lack of exploration is primarily due to the over-representation of women of European ancestry in genetic studies. Additionally, in contrast to earlier reports of a higher prevalence of ER- and TNBC in broadly defined “African” populations, more recent data show >50% ER+ breast cancer in some African populations (65), showing the critical need for more research into the prevalence and variation between populations in receptor status in breast cancers in African populations (66). We hope this data can help elucidate similarly powerful genetic predictors for women of East African descent. Only about 30% of heritable breast cancer can be attributed to known variants (7), and the majority of these are derived from patients with access to the best health care (8, 67).

## Materials and Methods

### Age of onset and population mortality rate analysis

Mortality rates for individuals from the age of 1-85+ years from a set of 100,000 women were downloaded from the World Health Organization (www.who.int). It is important to note that the mortality data from the American subset did not include any information about self-identified genetic background. These data were combined with breast cancer age of onset data from (1).

### Sample collection

We obtained tumor and matched normal tissue collected from 4 patients. From eligible participants, breast tissue specimens were collected within 10 minutes after surgery. The samples were soaked and preserved with phosphate buffered saline (PBS) pH 7 and subsequently transported within a cold chain to Tikur Anbessa Specialized Hospital and stored at -80 °C. All samples were deidentified by the Black Lion Hospital in Addis Ababa, Ethiopia, and no identifying information was shared with the authors of this study.

### Subtyping

Formalin fixed tissue samples were processed and paraffin-embedded, then subjected to immunohistochemistry for estrogen receptor at the Biorepository and Tissue Research core facility at The University of Virginia. The stains were interpreted by a Board-certified pathologist (C.A.M.). We pursued a subset of tumors that were ER^+^ for sequencing.

### Whole Exome Sequencing

Next-generation sequencing libraries were prepared using the Illumina TruSeq Exome kit with barcodes. Sequencing was done at the Genomics Core Facility at UMass Amherst on a NextSeq500. We used the TruSeq Exome kit (Illumina) to sequence matched normal and tumor tissue from 3 patients. Our data maps to hg38 at < 96%. Fastq files were mapped to the human reference genome (hg38) using Bowtie (68), and variants identified using mpileup in Samtools (69), and filtered using bcftools (70). Variants were called between the tumor-normal pairs using the VarScan2. Data were then compared with data generated by the TCGA Research Network: https://www.cancer.gov/tcga.

## Data Availability

Data is currently being archived and will be made available soon.

## Acknowledgements

We thank Trisha Zintel for her help with sequencing. We thank Carey Dougan and Ning-Hsuan Tseng for reading and commenting on this manuscript. This work was supported by a seed grant awarded to SRP and CCB from the Models 2 Medicine Center, part of the Institute for Applied Life Sciences at UMass Amherst. ADS was supported by a National Science Foundation Graduate Research Fellowship (Award 1451512). This work was supported by an NSF CAREER (DMR1454806), and NIH grants R21CA223783 and DP2CA186573 awarded to SRP. Funding was also provided in part by a generous donation from the Giglio Family to the Wallace H. Coulter Department of Biomedical Engineering (MOP). SRP was also supported by a grant from the Jane Koskinas Ted Giovanis Foundation for Health and Policy. SRP is an Armstrong Professional Development Professor.

