## Supplementary material for "Genetic Mutations Associated with Hormone-Positive Breast Cancer in Ethiopian Women": Table S1

| Ensembl ID | geneName | description |
|---|---|---|
| ENSG00000188984 | AADACL3 | arylacetamide deacetylase like 3 [Source:HGNC Symbol;Acc:HGNC:32037] |
| ENSG00000198691 | ABCA4 | ATP binding cassette subfamily A member 4 [Source:HGNC Symbol;Acc:HGNC:34] |
| ENSG00000117528 | ABCD3 | ATP binding cassette subfamily D member 3 [Source:HGNC Symbol;Acc:HGNC:67] |
| ENSG00000117054 | ACADM | acyl-CoA dehydrogenase medium chain [Source:HGNC Symbol;Acc:HGNC:89] |
| ENSG00000117054 | ACADM | acyl-CoA dehydrogenase medium chain [Source:HGNC Symbol;Acc:HGNC:89] |
| ENSG00000162390 | ACOT11 | acyl-CoA thioesterase 11 [Source:HGNC Symbol;Acc:HGNC:18156] |
| ENSG00000097021 | ACOT7 | acyl-CoA thioesterase 7 [Source:HGNC Symbol;Acc:HGNC:24157] |
| ENSG00000282608 | ADORA3 | adenosine A3 receptor [Source:HGNC Symbol;Acc:HGNC:268] |
| ENSG00000186094 | AGBL4 | ATP/GTP binding protein like 4 [Source:HGNC Symbol;Acc:HGNC:25892] |
| ENSG00000186094 | AGBL4 | ATP/GTP binding protein like 4 [Source:HGNC Symbol;Acc:HGNC:25892] |
| ENSG00000186094 | AGBL4 | ATP/GTP binding protein like 4 [Source:HGNC Symbol;Acc:HGNC:25892] |
| ENSG00000186094 | AGBL4 | ATP/GTP binding protein like 4 [Source:HGNC Symbol;Acc:HGNC:25892] |
| ENSG00000186094 | AGBL4 | ATP/GTP binding protein like 4 [Source:HGNC Symbol;Acc:HGNC:25892] |
| ENSG00000186094 | AGBL4 | ATP/GTP binding protein like 4 [Source:HGNC Symbol;Acc:HGNC:25892] |
| ENSG00000186094 | AGBL4 | ATP/GTP binding protein like 4 [Source:HGNC Symbol;Acc:HGNC:25892] |
| ENSG00000186094 | AGBL4 | ATP/GTP binding protein like 4 [Source:HGNC Symbol;Acc:HGNC:25892] |
| ENSG00000186094 | AGBL4 | ATP/GTP binding protein like 4 [Source:HGNC Symbol;Acc:HGNC:25892] |
| ENSG00000186094 | AGBL4 | ATP/GTP binding protein like 4 [Source:HGNC Symbol;Acc:HGNC:25892] |
| ENSG00000186094 | AGBL4 | ATP/GTP binding protein like 4 [Source:HGNC Symbol;Acc:HGNC:25892] |
| ENSG00000162688 | AGL | amylo-alpha-1, 6-glucosidase, 4-alpha-glucanotransferase [Source:HGNC Symbol;Acc:HGNC:321] |
| ENSG00000188157 | AGRN | agrin [Source:HGNC Symbol;Acc:HGNC:329] |
| ENSG00000188157 | AGRN | agrin [Source:HGNC Symbol;Acc:HGNC:329] |
| ENSG00000188157 | AGRN | agrin [Source:HGNC Symbol;Acc:HGNC:329] |
| ENSG00000196581 | AJAP1 | adherens junctions associated protein 1 [Source:HGNC Symbol;Acc:HGNC:30801] |
| ENSG00000162433 | AK4 | adenylate kinase 4 [Source:HGNC Symbol;Acc:HGNC:363] |
| ENSG00000154027 | AK5 | adenylate kinase 5 [Source:HGNC Symbol;Acc:HGNC:365] |
| ENSG00000154027 | AK5 | adenylate kinase 5 [Source:HGNC Symbol;Acc:HGNC:365] |
| ENSG00000154027 | AK5 | adenylate kinase 5 [Source:HGNC Symbol;Acc:HGNC:365] |
| ENSG00000174574 | AKIRIN1 | akirin 1 [Source:HGNC Symbol;Acc:HGNC:25744] |
| ENSG00000117448 | AKR1A1 | aldo-keto reductase family 1 member A1 [Source:HGNC Symbol;Acc:HGNC:380] |
| ENSG00000162482 | AKR7A3 | aldo-keto reductase family 7 member A3 [Source:HGNC Symbol;Acc:HGNC:390] |
| ENSG00000159423 | ALDH4A1 | aldehyde dehydrogenase 4 family member A1 [Source:HGNC Symbol;Acc:HGNC:406] |
| ENSG00000159423 | ALDH4A1 | aldehyde dehydrogenase 4 family member A1 [Source:HGNC Symbol;Acc:HGNC:406] |
| ENSG00000156150 | ALX3 | ALX homeobox 3 [Source:HGNC Symbol;Acc:HGNC:449] |
| ENSG00000074964 | ARHGEF10L | Rho guanine nucleotide exchange factor 10 like [Source:HGNC Symbol;Acc:HGNC:25540] |
| ENSG00000074964 | ARHGEF10L | Rho guanine nucleotide exchange factor 10 like [Source:HGNC Symbol;Acc:HGNC:25540] |
| ENSG00000074964 | ARHGEF10L | Rho guanine nucleotide exchange factor 10 like [Source:HGNC Symbol;Acc:HGNC:25540] |
| ENSG00000130762 | ARHGEF16 | Rho guanine nucleotide exchange factor 16 [Source:HGNC Symbol;Acc:HGNC:15515] |
| ENSG00000088280 | ASAP3 | ArfGAP with SH3 domain, ankyrin repeat and PH domain 3 [Source:HGNC Symbol;Acc:HGNC:14987] |
| ENSG00000088280 | ASAP3 | ArfGAP with SH3 domain, ankyrin repeat and PH domain 3 [Source:HGNC Symbol;Acc:HGNC:14987] |
| ENSG00000160072 | ATAD3B | ATPase family AAA domain containing 3B [Source:HGNC Symbol;Acc:HGNC:24007] |
| ENSG00000160072 | ATAD3B | ATPase family AAA domain containing 3B [Source:HGNC Symbol;Acc:HGNC:24007] |
| ENSG00000125703 | ATG4C | autophagy related 4C cysteine peptidase [Source:HGNC Symbol;Acc:HGNC:16040] |
| ENSG00000123472 | ATPAF1 | ATP synthase mitochondrial F1 complex assembly factor 1 [Source:HGNC Symbol;Acc:HGNC:18803] |
| ENSG00000123472 | ATPAF1 | ATP synthase mitochondrial F1 complex assembly factor 1 [Source:HGNC Symbol;Acc:HGNC:18803] |
| ENSG00000137936 | BCAR3 | BCAR3 adaptor protein, NSP family member [Source:HGNC Symbol;Acc:HGNC:973] |
| ENSG00000137936 | BCAR3 | BCAR3 adaptor protein, NSP family member [Source:HGNC Symbol;Acc:HGNC:973] |
| ENSG00000183682 | BMP8A | bone morphogenetic protein 8a [Source:HGNC Symbol;Acc:HGNC:21650] |
| ENSG00000183682 | BMP8A | bone morphogenetic protein 8a [Source:HGNC Symbol;Acc:HGNC:21650] |
| ENSG00000137948 | BRDT | bromodomain testis associated [Source:HGNC Symbol;Acc:HGNC:1105] |
| None | None | None |
| None | None | None |
| None | None | None |
| ENSG00000162598 | C1orf87 | chromosome 1 open reading frame 87 [Source:HGNC Symbol;Acc:HGNC:28547] |
| ENSG00000158966 | CACHD1 | cache domain containing 1 [Source:HGNC Symbol;Acc:HGNC:29314] |
| ENSG00000158966 | CACHD1 | cache domain containing 1 [Source:HGNC Symbol;Acc:HGNC:29314] |
| ENSG00000171735 | CAMTA1 | calmodulin binding transcription activator 1 [Source:HGNC Symbol;Acc:HGNC:18806] |
| ENSG00000171735 | CAMTA1 | calmodulin binding transcription activator 1 [Source:HGNC Symbol;Acc:HGNC:18806] |
| ENSG00000171735 | CAMTA1 | calmodulin binding transcription activator 1 [Source:HGNC Symbol;Acc:HGNC:18806] |
| ENSG00000171735 | CAMTA1 | calmodulin binding transcription activator 1 [Source:HGNC Symbol;Acc:HGNC:18806] |
| ENSG00000171735 | CAMTA1 | calmodulin binding transcription activator 1 [Source:HGNC Symbol;Acc:HGNC:18806] |
| ENSG00000171735 | CAMTA1 | calmodulin binding transcription activator 1 [Source:HGNC Symbol;Acc:HGNC:18806] |
| ENSG00000171735 | CAMTA1 | calmodulin binding transcription activator 1 [Source:HGNC Symbol;Acc:HGNC:18806] |
| ENSG00000171735 | CAMTA1 | calmodulin binding transcription activator 1 [Source:HGNC Symbol;Acc:HGNC:18806] |
| ENSG00000171735 | CAMTA1 | calmodulin binding transcription activator 1 [Source:HGNC Symbol;Acc:HGNC:18806] |
| ENSG00000171735 | CAMTA1 | calmodulin binding transcription activator 1 [Source:HGNC Symbol;Acc:HGNC:18806] |
| ENSG00000116489 | CAPZA1 | capping actin protein of muscle Z-line subunit alpha 1 [Source:HGNC Symbol;Acc:HGNC:1488] |
| ENSG00000077549 | CAPZB | capping actin protein of muscle Z-line subunit beta [Source:HGNC Symbol;Acc:HGNC:1491] |
| ENSG00000132906 | CASP9 | caspase 9 [Source:HGNC Symbol;Acc:HGNC:1511] |
| ENSG00000118729 | CASQ2 | calsequestrin 2 [Source:HGNC Symbol;Acc:HGNC:1513] |
| ENSG00000130940 | CASZ1 | castor zinc finger 1 [Source:HGNC Symbol;Acc:HGNC:26002] |
| ENSG00000130940 | CASZ1 | castor zinc finger 1 [Source:HGNC Symbol;Acc:HGNC:26002] |
| ENSG00000188782 | CATSPER4 | cation channel sperm associated 4 [Source:HGNC Symbol;Acc:HGNC:23220] |
| ENSG00000122483 | CCDC18 | coiled-coil domain containing 18 [Source:HGNC Symbol;Acc:HGNC:30370] |
| ENSG00000122483 | CCDC18 | coiled-coil domain containing 18 [Source:HGNC Symbol;Acc:HGNC:30370] |
| ENSG00000122483 | CCDC18 | coiled-coil domain containing 18 [Source:HGNC Symbol;Acc:HGNC:30370] |
| ENSG00000162592 | CCDC27 | coiled-coil domain containing 27 [Source:HGNC Symbol;Acc:HGNC:26546] |

| | | |
|---|---|---|
| ENSG00000186409 | CCDC30 | coiled-coil domain containing 30 [Source:HGNC Symbol;Acc:HGNC:26103] |
| ENSG00000186409 | CCDC30 | coiled-coil domain containing 30 [Source:HGNC Symbol;Acc:HGNC:26103] |
| ENSG00000079335 | CDC14A | cell division cycle 14A [Source:HGNC Symbol;Acc:HGNC:1718] |
| ENSG00000157211 | CDCP2 | CUB domain containing protein 2 [Source:HGNC Symbol;Acc:HGNC:27297] |
| ENSG00000157211 | CDCP2 | CUB domain containing protein 2 [Source:HGNC Symbol;Acc:HGNC:27297] |
| None | None | None |
| None | None | None |
| ENSG00000248333 | CDK11B | cyclin dependent kinase 11B [Source:HGNC Symbol;Acc:HGNC:1729] |
| ENSG00000142615 | CELA2A | chymotrypsin like elastase 2A [Source:HGNC Symbol;Acc:HGNC:24609] |
| ENSG00000215704 | CELA2B | chymotrypsin like elastase 2B [Source:HGNC Symbol;Acc:HGNC:29995] |
| ENSG00000219073 | CELA3B | chymotrypsin like elastase 3B [Source:HGNC Symbol;Acc:HGNC:15945] |
| ENSG00000219073 | CELA3B | chymotrypsin like elastase 3B [Source:HGNC Symbol;Acc:HGNC:15945] |
| ENSG00000116198 | CEP104 | centrosomal protein 104 [Source:HGNC Symbol;Acc:HGNC:24866] |
| ENSG00000116198 | CEP104 | centrosomal protein 104 [Source:HGNC Symbol;Acc:HGNC:24866] |
| ENSG00000130695 | CEP85 | centrosomal protein 85 [Source:HGNC Symbol;Acc:HGNC:25309] |
| ENSG00000016490 | CLCA1 | chloride channel accessory 1 [Source:HGNC Symbol;Acc:HGNC:2015] |
| ENSG00000011021 | CLCN6 | chloride voltage-gated channel 6 [Source:HGNC Symbol;Acc:HGNC:2024] |
| ENSG00000011021 | CLCN6 | chloride voltage-gated channel 6 [Source:HGNC Symbol;Acc:HGNC:2024] |
| ENSG00000186510 | CLCNKA | chloride voltage-gated channel Ka [Source:HGNC Symbol;Acc:HGNC:2026] |
| ENSG00000092853 | CLSPN | claspin [Source:HGNC Symbol;Acc:HGNC:19715] |
| ENSG00000188822 | CNR2 | cannabinoid receptor 2 [Source:HGNC Symbol;Acc:HGNC:2160] |
| ENSG00000060718 | COL11A1 | collagen type XI alpha 1 chain [Source:HGNC Symbol;Acc:HGNC:2186] |
| ENSG00000060718 | COL11A1 | collagen type XI alpha 1 chain [Source:HGNC Symbol;Acc:HGNC:2186] |
| ENSG00000060718 | COL11A1 | collagen type XI alpha 1 chain [Source:HGNC Symbol;Acc:HGNC:2186] |
| ENSG00000060718 | COL11A1 | collagen type XI alpha 1 chain [Source:HGNC Symbol;Acc:HGNC:2186] |
| ENSG00000084636 | COL16A1 | collagen type XVI alpha 1 chain [Source:HGNC Symbol;Acc:HGNC:2193] |
| ENSG00000171502 | COL24A1 | collagen type XXIV alpha 1 chain [Source:HGNC Symbol;Acc:HGNC:20821] |
| ENSG00000171502 | COL24A1 | collagen type XXIV alpha 1 chain [Source:HGNC Symbol;Acc:HGNC:20821] |
| ENSG00000171502 | COL24A1 | collagen type XXIV alpha 1 chain [Source:HGNC Symbol;Acc:HGNC:20821] |
| ENSG00000058453 | CROCC | ciliary rootlet coiled-coil, rootletin [Source:HGNC Symbol;Acc:HGNC:21299] |
| ENSG00000058453 | CROCC | ciliary rootlet coiled-coil, rootletin [Source:HGNC Symbol;Acc:HGNC:21299] |
| ENSG00000058453 | CROCC | ciliary rootlet coiled-coil, rootletin [Source:HGNC Symbol;Acc:HGNC:21299] |
| ENSG00000215908 | CROCCP2 | CROCC pseudogene 2 [Source:HGNC Symbol;Acc:HGNC:28170] |
| ENSG00000215908 | CROCCP2 | CROCC pseudogene 2 [Source:HGNC Symbol;Acc:HGNC:28170] |
| ENSG00000215908 | CROCCP2 | CROCC pseudogene 2 [Source:HGNC Symbol;Acc:HGNC:28170] |
| ENSG00000215908 | CROCCP2 | CROCC pseudogene 2 [Source:HGNC Symbol;Acc:HGNC:28170] |
| ENSG00000080947 | CROCCP3 | CROCC pseudogene 3 [Source:HGNC Symbol;Acc:HGNC:29405] |
| ENSG00000121904 | CSMD2 | CUB and Sushi multiple domains 2 [Source:HGNC Symbol;Acc:HGNC:19290] |
| ENSG00000121904 | CSMD2 | CUB and Sushi multiple domains 2 [Source:HGNC Symbol;Acc:HGNC:19290] |
| ENSG00000121904 | CSMD2 | CUB and Sushi multiple domains 2 [Source:HGNC Symbol;Acc:HGNC:19290] |
| ENSG00000121904 | CSMD2 | CUB and Sushi multiple domains 2 [Source:HGNC Symbol;Acc:HGNC:19290] |
| ENSG00000121904 | CSMD2 | CUB and Sushi multiple domains 2 [Source:HGNC Symbol;Acc:HGNC:19290] |
| ENSG00000121904 | CSMD2 | CUB and Sushi multiple domains 2 [Source:HGNC Symbol;Acc:HGNC:19290] |
| ENSG00000143079 | CTTNBP2NL | CTTNBP2 N-terminal like [Source:HGNC Symbol;Acc:HGNC:25330] |
| ENSG00000143079 | CTTNBP2NL | CTTNBP2 N-terminal like [Source:HGNC Symbol;Acc:HGNC:25330] |
| ENSG00000143079 | CTTNBP2NL | CTTNBP2 N-terminal like [Source:HGNC Symbol;Acc:HGNC:25330] |
| ENSG00000174151 | CYB561D1 | cytochrome b561 family member D1 [Source:HGNC Symbol;Acc:HGNC:26804] |
| ENSG00000154198 | CYP4Z2P | cytochrome P450 family 4 subfamily Z member 2, pseudogene [Source:HGNC Symbol;Acc:HGNC:24426] |
| ENSG00000173406 | DAB1 | DAB adaptor protein 1 [Source:HGNC Symbol;Acc:HGNC:2661] |
| ENSG00000173406 | DAB1 | DAB adaptor protein 1 [Source:HGNC Symbol;Acc:HGNC:2661] |
| ENSG00000173406 | DAB1 | DAB adaptor protein 1 [Source:HGNC Symbol;Acc:HGNC:2661] |
| ENSG00000173406 | DAB1 | DAB adaptor protein 1 [Source:HGNC Symbol;Acc:HGNC:2661] |
| ENSG00000173406 | DAB1 | DAB adaptor protein 1 [Source:HGNC Symbol;Acc:HGNC:2661] |
| ENSG00000173406 | DAB1 | DAB adaptor protein 1 [Source:HGNC Symbol;Acc:HGNC:2661] |
| ENSG00000173406 | DAB1 | DAB adaptor protein 1 [Source:HGNC Symbol;Acc:HGNC:2661] |
| ENSG00000173406 | DAB1 | DAB adaptor protein 1 [Source:HGNC Symbol;Acc:HGNC:2661] |
| ENSG00000173406 | DAB1 | DAB adaptor protein 1 [Source:HGNC Symbol;Acc:HGNC:2661] |
| ENSG00000173406 | DAB1 | DAB adaptor protein 1 [Source:HGNC Symbol;Acc:HGNC:2661] |
| ENSG00000173406 | DAB1 | DAB adaptor protein 1 [Source:HGNC Symbol;Acc:HGNC:2661] |
| ENSG00000173406 | DAB1 | DAB adaptor protein 1 [Source:HGNC Symbol;Acc:HGNC:2661] |
| ENSG00000173406 | DAB1 | DAB adaptor protein 1 [Source:HGNC Symbol;Acc:HGNC:2661] |
| ENSG00000153904 | DDAH1 | dimethylarginine dimethylaminohydrolase 1 [Source:HGNC Symbol;Acc:HGNC:2715] |
| ENSG00000153904 | DDAH1 | dimethylarginine dimethylaminohydrolase 1 [Source:HGNC Symbol;Acc:HGNC:2715] |
| ENSG00000153904 | DDAH1 | dimethylarginine dimethylaminohydrolase 1 [Source:HGNC Symbol;Acc:HGNC:2715] |
| ENSG00000153904 | DDAH1 | dimethylarginine dimethylaminohydrolase 1 [Source:HGNC Symbol;Acc:HGNC:2715] |
| ENSG00000197312 | DDI2 | DNA damage inducible 1 homolog 2 [Source:HGNC Symbol;Acc:HGNC:24578] |
| ENSG00000175984 | DENND2C | DENN domain containing 2C [Source:HGNC Symbol;Acc:HGNC:24748] |
| ENSG00000175984 | DENND2C | DENN domain containing 2C [Source:HGNC Symbol;Acc:HGNC:24748] |
| ENSG00000169598 | DFFB | DNA fragmentation factor subunit beta [Source:HGNC Symbol;Acc:HGNC:2773] |
| ENSG00000169598 | DFFB | DNA fragmentation factor subunit beta [Source:HGNC Symbol;Acc:HGNC:2773] |
| ENSG00000169598 | DFFB | DNA fragmentation factor subunit beta [Source:HGNC Symbol;Acc:HGNC:2773] |
| ENSG00000116133 | DHCR24 | 24-dehydrocholesterol reductase [Source:HGNC Symbol;Acc:HGNC:2859] |
| ENSG00000007923 | DNAJC11 | DnaJ heat shock protein family (Hsp40) member C11 [Source:HGNC Symbol;Acc:HGNC:25570] |
| ENSG00000116675 | DNAJC6 | DnaJ heat shock protein family (Hsp40) member C6 [Source:HGNC Symbol;Acc:HGNC:15469] |
| ENSG00000116675 | DNAJC6 | DnaJ heat shock protein family (Hsp40) member C6 [Source:HGNC Symbol;Acc:HGNC:15469] |
| ENSG00000116675 | DNAJC6 | DnaJ heat shock protein family (Hsp40) member C6 [Source:HGNC Symbol;Acc:HGNC:15469] |
| ENSG00000116675 | DNAJC6 | DnaJ heat shock protein family (Hsp40) member C6 [Source:HGNC Symbol;Acc:HGNC:15469] |
| ENSG00000116675 | DNAJC6 | DnaJ heat shock protein family (Hsp40) member C6 [Source:HGNC Symbol;Acc:HGNC:15469] |

| | | |
|---|---|---|
| ENSG00000116641 | DOCK7 | dedicator of cytokinesis 7 [Source:HGNC Symbol;Acc:HGNC:19190] |
| ENSG00000116641 | DOCK7 | dedicator of cytokinesis 7 [Source:HGNC Symbol;Acc:HGNC:19190] |
| ENSG00000116641 | DOCK7 | dedicator of cytokinesis 7 [Source:HGNC Symbol;Acc:HGNC:19190] |
| ENSG00000188641 | DPYD | dihydropyrimidine dehydrogenase [Source:HGNC Symbol;Acc:HGNC:3012] |
| ENSG00000232878 | DPYD-AS1 | DPYD antisense RNA 1 [Source:HGNC Symbol;Acc:HGNC:40195] |
| ENSG00000232878 | DPYD-AS1 | DPYD antisense RNA 1 [Source:HGNC Symbol;Acc:HGNC:40195] |
| ENSG00000117298 | ECE1 | endothelin converting enzyme 1 [Source:HGNC Symbol;Acc:HGNC:3146] |
| ENSG00000203965 | EFCAB7 | EF-hand calcium binding domain 7 [Source:HGNC Symbol;Acc:HGNC:29379] |
| ENSG00000142634 | EFHD2 | EF-hand domain family member D2 [Source:HGNC Symbol;Acc:HGNC:28670] |
| ENSG00000070785 | EIF2B3 | eukaryotic translation initiation factor 2B subunit gamma [Source:HGNC Symbol;Acc:HGNC:3259] |
| ENSG00000075151 | EIF4G3 | eukaryotic translation initiation factor 4 gamma 3 [Source:HGNC Symbol;Acc:HGNC:3298] |
| ENSG00000075151 | EIF4G3 | eukaryotic translation initiation factor 4 gamma 3 [Source:HGNC Symbol;Acc:HGNC:3298] |
| ENSG00000075151 | EIF4G3 | eukaryotic translation initiation factor 4 gamma 3 [Source:HGNC Symbol;Acc:HGNC:3298] |
| ENSG00000075151 | EIF4G3 | eukaryotic translation initiation factor 4 gamma 3 [Source:HGNC Symbol;Acc:HGNC:3298] |
| None | None | None |
| None | None | None |
| ENSG00000159023 | EPB41 | erythrocyte membrane protein band 4.1 [Source:HGNC Symbol;Acc:HGNC:3377] |
| ENSG00000183317 | EPHA10 | EPH receptor A10 [Source:HGNC Symbol;Acc:HGNC:19987] |
| ENSG00000070886 | EPHA8 | EPH receptor A8 [Source:HGNC Symbol;Acc:HGNC:3391] |
| ENSG00000070886 | EPHA8 | EPH receptor A8 [Source:HGNC Symbol;Acc:HGNC:3391] |
| ENSG00000133216 | EPHB2 | EPH receptor B2 [Source:HGNC Symbol;Acc:HGNC:3393] |
| ENSG00000198758 | EPS8L3 | EPS8 like 3 [Source:HGNC Symbol;Acc:HGNC:21297] |
| ENSG00000117419 | ERI3 | ERI1 exoribonuclease family member 3 [Source:HGNC Symbol;Acc:HGNC:17276] |
| ENSG00000268869 | ESPNP | espin pseudogene [Source:HGNC Symbol;Acc:HGNC:23285] |
| ENSG00000268869 | ESPNP | espin pseudogene [Source:HGNC Symbol;Acc:HGNC:23285] |
| ENSG00000268869 | ESPNP | espin pseudogene [Source:HGNC Symbol;Acc:HGNC:23285] |
| ENSG00000067208 | EVI5 | ecotropic viral integration site 5 [Source:HGNC Symbol;Acc:HGNC:3501] |
| ENSG00000067208 | EVI5 | ecotropic viral integration site 5 [Source:HGNC Symbol;Acc:HGNC:3501] |
| ENSG00000067208 | EVI5 | ecotropic viral integration site 5 [Source:HGNC Symbol;Acc:HGNC:3501] |
| ENSG00000067208 | EVI5 | ecotropic viral integration site 5 [Source:HGNC Symbol;Acc:HGNC:3501] |
| ENSG00000185104 | FAF1 | Fas associated factor 1 [Source:HGNC Symbol;Acc:HGNC:3578] |
| ENSG00000185104 | FAF1 | Fas associated factor 1 [Source:HGNC Symbol;Acc:HGNC:3578] |
| ENSG00000185104 | FAF1 | Fas associated factor 1 [Source:HGNC Symbol;Acc:HGNC:3578] |
| ENSG00000172456 | FGGY | FGGY carbohydrate kinase domain containing [Source:HGNC Symbol;Acc:HGNC:25610] |
| ENSG00000172456 | FGGY | FGGY carbohydrate kinase domain containing [Source:HGNC Symbol;Acc:HGNC:25610] |
| ENSG00000142621 | FHAD1 | forkhead associated phosphopeptide binding domain 1 [Source:HGNC Symbol;Acc:HGNC:29408] |
| ENSG00000142621 | FHAD1 | forkhead associated phosphopeptide binding domain 1 [Source:HGNC Symbol;Acc:HGNC:29408] |
| ENSG00000142621 | FHAD1 | forkhead associated phosphopeptide binding domain 1 [Source:HGNC Symbol;Acc:HGNC:29408] |
| ENSG00000142621 | FHAD1 | forkhead associated phosphopeptide binding domain 1 [Source:HGNC Symbol;Acc:HGNC:29408] |
| ENSG00000142621 | FHAD1 | forkhead associated phosphopeptide binding domain 1 [Source:HGNC Symbol;Acc:HGNC:29408] |
| ENSG00000198815 | FOXJ3 | forkhead box J3 [Source:HGNC Symbol;Acc:HGNC:29178] |
| ENSG00000198815 | FOXJ3 | forkhead box J3 [Source:HGNC Symbol;Acc:HGNC:29178] |
| None | None | None |
| None | None | None |
| None | None | None |
| None | None | None |
| ENSG00000156869 | FRRS1 | ferric chelate reductase 1 [Source:HGNC Symbol;Acc:HGNC:27622] |
| ENSG00000183347 | GBP6 | guanylate binding protein family member 6 [Source:HGNC Symbol;Acc:HGNC:25395] |
| ENSG00000213512 | GBP7 | guanylate binding protein 7 [Source:HGNC Symbol;Acc:HGNC:29606] |
| ENSG00000137960 | GIPC2 | GIPC PDZ domain containing family member 2 [Source:HGNC Symbol;Acc:HGNC:18177] |
| ENSG00000174332 | GLIS1 | GLIS family zinc finger 1 [Source:HGNC Symbol;Acc:HGNC:29525] |
| ENSG00000174332 | GLIS1 | GLIS family zinc finger 1 [Source:HGNC Symbol;Acc:HGNC:29525] |
| ENSG00000174332 | GLIS1 | GLIS family zinc finger 1 [Source:HGNC Symbol;Acc:HGNC:29525] |
| ENSG00000174332 | GLIS1 | GLIS family zinc finger 1 [Source:HGNC Symbol;Acc:HGNC:29525] |
| ENSG00000174332 | GLIS1 | GLIS family zinc finger 1 [Source:HGNC Symbol;Acc:HGNC:29525] |
| ENSG00000078369 | GNB1 | G protein subunit beta 1 [Source:HGNC Symbol;Acc:HGNC:4396] |
| ENSG00000172380 | GNG12 | G protein subunit gamma 12 [Source:HGNC Symbol;Acc:HGNC:19663] |
| ENSG00000232284 | GNG12-AS1 | GNG12, DIRAS3 and WLS antisense RNA 1 [Source:HGNC Symbol;Acc:HGNC:43938] |
| ENSG00000232284 | GNG12-AS1 | GNG12, DIRAS3 and WLS antisense RNA 1 [Source:HGNC Symbol;Acc:HGNC:43938] |
| ENSG00000232284 | GNG12-AS1 | GNG12, DIRAS3 and WLS antisense RNA 1 [Source:HGNC Symbol;Acc:HGNC:43938] |
| ENSG00000159592 | GPBP1L1 | GC-rich promoter binding protein 1 like 1 [Source:HGNC Symbol;Acc:HGNC:28843] |
| ENSG00000158055 | GRHL3 | grainyhead like transcription factor 3 [Source:HGNC Symbol;Acc:HGNC:25839] |
| ENSG00000163873 | GRIK3 | glutamate ionotropic receptor kainate type subunit 3 [Source:HGNC Symbol;Acc:HGNC:4581] |
| ENSG00000163873 | GRIK3 | glutamate ionotropic receptor kainate type subunit 3 [Source:HGNC Symbol;Acc:HGNC:4581] |
| ENSG00000162669 | HFM1 | helicase for meiosis 1 [Source:HGNC Symbol;Acc:HGNC:20193] |
| ENSG00000162669 | HFM1 | helicase for meiosis 1 [Source:HGNC Symbol;Acc:HGNC:20193] |
| ENSG00000162669 | HFM1 | helicase for meiosis 1 [Source:HGNC Symbol;Acc:HGNC:20193] |
| ENSG00000127124 | HIVEP3 | HIVEP zinc finger 3 [Source:HGNC Symbol;Acc:HGNC:13561] |
| ENSG00000127124 | HIVEP3 | HIVEP zinc finger 3 [Source:HGNC Symbol;Acc:HGNC:13561] |
| ENSG00000127124 | HIVEP3 | HIVEP zinc finger 3 [Source:HGNC Symbol;Acc:HGNC:13561] |
| ENSG00000127124 | HIVEP3 | HIVEP zinc finger 3 [Source:HGNC Symbol;Acc:HGNC:13561] |
| ENSG00000127124 | HIVEP3 | HIVEP zinc finger 3 [Source:HGNC Symbol;Acc:HGNC:13561] |
| ENSG00000127124 | HIVEP3 | HIVEP zinc finger 3 [Source:HGNC Symbol;Acc:HGNC:13561] |
| ENSG00000153936 | HS2ST1 | heparan sulfate 2-O-sulfotransferase 1 [Source:HGNC Symbol;Acc:HGNC:5193] |
| ENSG00000153936 | HS2ST1 | heparan sulfate 2-O-sulfotransferase 1 [Source:HGNC Symbol;Acc:HGNC:5193] |
| ENSG00000136720 | HS6ST1 | heparan sulfate 6-O-sulfotransferase 1 [Source:HGNC Symbol;Acc:HGNC:5201] |
| ENSG00000136720 | HS6ST1 | heparan sulfate 6-O-sulfotransferase 1 [Source:HGNC Symbol;Acc:HGNC:5201] |
| ENSG00000136720 | HS6ST1 | heparan sulfate 6-O-sulfotransferase 1 [Source:HGNC Symbol;Acc:HGNC:5201] |

| | | |
|---|---|---|
| ENSG00000142798 | HSPG2 | heparan sulfate proteoglycan 2 [Source:HGNC Symbol;Acc:HGNC:5273] |
| ENSG00000142798 | HSPG2 | heparan sulfate proteoglycan 2 [Source:HGNC Symbol;Acc:HGNC:5273] |
| ENSG00000142798 | HSPG2 | heparan sulfate proteoglycan 2 [Source:HGNC Symbol;Acc:HGNC:5273] |
| ENSG00000142798 | HSPG2 | heparan sulfate proteoglycan 2 [Source:HGNC Symbol;Acc:HGNC:5273] |
| ENSG00000185436 | IFNLR1 | interferon lambda receptor 1 [Source:HGNC Symbol;Acc:HGNC:18584] |
| ENSG00000117154 | IGSF21 | immunoglobin superfamily member 21 [Source:HGNC Symbol;Acc:HGNC:28246] |
| ENSG00000117154 | IGSF21 | immunoglobin superfamily member 21 [Source:HGNC Symbol;Acc:HGNC:28246] |
| ENSG00000117154 | IGSF21 | immunoglobin superfamily member 21 [Source:HGNC Symbol;Acc:HGNC:28246] |
| ENSG00000117154 | IGSF21 | immunoglobin superfamily member 21 [Source:HGNC Symbol;Acc:HGNC:28246] |
| ENSG00000081985 | IL12RB2 | interleukin 12 receptor subunit beta 2 [Source:HGNC Symbol;Acc:HGNC:5972] |
| ENSG00000081985 | IL12RB2 | interleukin 12 receptor subunit beta 2 [Source:HGNC Symbol;Acc:HGNC:5972] |
| ENSG00000142677 | IL22RA1 | interleukin 22 receptor subunit alpha 1 [Source:HGNC Symbol;Acc:HGNC:13700] |
| ENSG00000162594 | IL23R | interleukin 23 receptor [Source:HGNC Symbol;Acc:HGNC:19100] |
| None | None | None |
| None | None | None |
| None | None | None |
| None | None | None |
| None | None | None |
| ENSG00000204084 | INPP5B | inositol polyphosphate-5-phosphatase B [Source:HGNC Symbol;Acc:HGNC:6077] |
| ENSG00000204084 | INPP5B | inositol polyphosphate-5-phosphatase B [Source:HGNC Symbol;Acc:HGNC:6077] |
| ENSG00000197429 | IPP | intracisternal A particle-promoted polypeptide [Source:HGNC Symbol;Acc:HGNC:6108] |
| ENSG00000142856 | ITGB3BP | integrin subunit beta 3 binding protein [Source:HGNC Symbol;Acc:HGNC:6157] |
| ENSG00000162434 | JAK1 | Janus kinase 1 [Source:HGNC Symbol;Acc:HGNC:6190] |
| ENSG00000162434 | JAK1 | Janus kinase 1 [Source:HGNC Symbol;Acc:HGNC:6190] |
| ENSG00000132854 | KANK4 | KN motif and ankyrin repeat domains 4 [Source:HGNC Symbol;Acc:HGNC:27263] |
| ENSG00000132854 | KANK4 | KN motif and ankyrin repeat domains 4 [Source:HGNC Symbol;Acc:HGNC:27263] |
| ENSG00000189337 | KAZN | kazrin, periplakin interacting protein [Source:HGNC Symbol;Acc:HGNC:29173] |
| ENSG00000189337 | KAZN | kazrin, periplakin interacting protein [Source:HGNC Symbol;Acc:HGNC:29173] |
| ENSG00000189337 | KAZN | kazrin, periplakin interacting protein [Source:HGNC Symbol;Acc:HGNC:29173] |
| ENSG00000189337 | KAZN | kazrin, periplakin interacting protein [Source:HGNC Symbol;Acc:HGNC:29173] |
| ENSG00000189337 | KAZN | kazrin, periplakin interacting protein [Source:HGNC Symbol;Acc:HGNC:29173] |
| ENSG00000189337 | KAZN | kazrin, periplakin interacting protein [Source:HGNC Symbol;Acc:HGNC:29173] |
| ENSG00000189337 | KAZN | kazrin, periplakin interacting protein [Source:HGNC Symbol;Acc:HGNC:29173] |
| ENSG00000069424 | KCNAB2 | potassium voltage-gated channel subfamily A regulatory beta subunit 2 [Source:HGNC Symbol;Acc:HGNC:6229] |
| ENSG00000069424 | KCNAB2 | potassium voltage-gated channel subfamily A regulatory beta subunit 2 [Source:HGNC Symbol;Acc:HGNC:6229] |
| ENSG00000116396 | KCNC4 | potassium voltage-gated channel subfamily C member 4 [Source:HGNC Symbol;Acc:HGNC:6236] |
| ENSG00000171385 | KCND3 | potassium voltage-gated channel subfamily D member 3 [Source:HGNC Symbol;Acc:HGNC:6239] |
| ENSG00000117013 | KCNQ4 | potassium voltage-gated channel subfamily Q member 4 [Source:HGNC Symbol;Acc:HGNC:6298] |
| ENSG00000004487 | KDM1A | lysine demethylase 1A [Source:HGNC Symbol;Acc:HGNC:29079] |
| ENSG00000116299 | KIAA1324 | KIAA1324 [Source:HGNC Symbol;Acc:HGNC:29618] |
| ENSG00000162522 | KIAA1522 | KIAA1522 [Source:HGNC Symbol;Acc:HGNC:29301] |
| ENSG00000162522 | KIAA1522 | KIAA1522 [Source:HGNC Symbol;Acc:HGNC:29301] |
| None | None | None |
| None | None | None |
| ENSG00000117245 | KIF17 | kinesin family member 17 [Source:HGNC Symbol;Acc:HGNC:19167] |
| ENSG00000117245 | KIF17 | kinesin family member 17 [Source:HGNC Symbol;Acc:HGNC:19167] |
| ENSG00000142945 | KIF2C | kinesin family member 2C [Source:HGNC Symbol;Acc:HGNC:6393] |
| ENSG00000055609 | KMT2C | lysine methyltransferase 2C [Source:HGNC Symbol;Acc:HGNC:13726] |
| ENSG00000162511 | LAPTM5 | lysosomal protein transmembrane 5 [Source:HGNC Symbol;Acc:HGNC:29612] |
| ENSG00000162511 | LAPTM5 | lysosomal protein transmembrane 5 [Source:HGNC Symbol;Acc:HGNC:29612] |
| ENSG00000116678 | LEPR | leptin receptor [Source:HGNC Symbol;Acc:HGNC:6554] |
| ENSG00000116678 | LEPR | leptin receptor [Source:HGNC Symbol;Acc:HGNC:6554] |
| ENSG00000116678 | LEPR | leptin receptor [Source:HGNC Symbol;Acc:HGNC:6554] |
| ENSG00000224209 | LINC00466 | long intergenic non-protein coding RNA 466 [Source:HGNC Symbol;Acc:HGNC:27294] |
| ENSG00000224209 | LINC00466 | long intergenic non-protein coding RNA 466 [Source:HGNC Symbol;Acc:HGNC:27294] |
| None | None | None |
| None | None | None |
| None | None | None |
| None | None | None |
| None | None | None |
| None | None | None |
| None | None | None |
| None | None | None |
| None | None | None |
| None | None | None |
| None | None | None |
| None | None | None |
| None | None | None |
| None | None | None |
| None | None | None |
| None | None | None |
| None | None | None |
| None | None | None |
| None | None | None |
| None | None | None |
| None | None | None |
| None | None | None |

| | | |
|---|---|---|
| None | None | None |
| None | None | None |
| None | None | None |
| None | None | None |
| None | None | None |
| None | None | None |
| None | None | None |
| None | None | None |
| None | None | None |
| None | None | None |
| None | None | None |
| None | None | None |
| None | None | None |
| None | None | None |
| None | None | None |
| None | None | None |
| None | None | None |
| None | None | None |
| None | None | None |
| None | None | None |
| None | None | None |
| None | None | None |
| None | None | None |
| None | None | None |
| None | None | None |
| None | None | None |
| None | None | None |
| None | None | None |
| None | None | None |
| None | None | None |
| None | None | None |
| None | None | None |
| None | None | None |
| None | None | None |
| None | None | None |
| None | None | None |
| ENSG00000157193 | LRP8 | LDL receptor related protein 8 [Source:HGNC Symbol;Acc:HGNC:6700] |
| ENSG00000157193 | LRP8 | LDL receptor related protein 8 [Source:HGNC Symbol;Acc:HGNC:6700] |
| ENSG00000162494 | LRRC38 | leucine rich repeat containing 38 [Source:HGNC Symbol;Acc:HGNC:27005] |
| ENSG00000130764 | LRRC47 | leucine rich repeat containing 47 [Source:HGNC Symbol;Acc:HGNC:29207] |
| ENSG00000033122 | LRRC7 | leucine rich repeat containing 7 [Source:HGNC Symbol;Acc:HGNC:18531] |
| ENSG00000162620 | LRRIQ3 | leucine rich repeats and IQ motif containing 3 [Source:HGNC Symbol;Acc:HGNC:28318] |
| ENSG00000127603 | MACF1 | microtubule actin crosslinking factor 1 [Source:HGNC Symbol;Acc:HGNC:13664] |
| ENSG00000127603 | MACF1 | microtubule actin crosslinking factor 1 [Source:HGNC Symbol;Acc:HGNC:13664] |
| ENSG00000127603 | MACF1 | microtubule actin crosslinking factor 1 [Source:HGNC Symbol;Acc:HGNC:13664] |
| ENSG00000127603 | MACF1 | microtubule actin crosslinking factor 1 [Source:HGNC Symbol;Acc:HGNC:13664] |
| ENSG00000081026 | MAGI3 | membrane associated guanylate kinase, WW and PDZ domain containing 3 [Source:HGNC Symbol;Acc:HGNC:29647] |
| ENSG00000081026 | MAGI3 | membrane associated guanylate kinase, WW and PDZ domain containing 3 [Source:HGNC Symbol;Acc:HGNC:29647] |
| ENSG00000081026 | MAGI3 | membrane associated guanylate kinase, WW and PDZ domain containing 3 [Source:HGNC Symbol;Acc:HGNC:29647] |
| ENSG00000117643 | MAN1C1 | mannosidase alpha class 1C member 1 [Source:HGNC Symbol;Acc:HGNC:19080] |
| ENSG00000117643 | MAN1C1 | mannosidase alpha class 1C member 1 [Source:HGNC Symbol;Acc:HGNC:19080] |
| ENSG00000117643 | MAN1C1 | mannosidase alpha class 1C member 1 [Source:HGNC Symbol;Acc:HGNC:19080] |
| ENSG00000117643 | MAN1C1 | mannosidase alpha class 1C member 1 [Source:HGNC Symbol;Acc:HGNC:19080] |
| ENSG00000116871 | MAP7D1 | MAP7 domain containing 1 [Source:HGNC Symbol;Acc:HGNC:25514] |
| ENSG00000009724 | MASP2 | mannan binding lectin serine peptidase 2 [Source:HGNC Symbol;Acc:HGNC:6902] |
| ENSG00000086015 | MAST2 | microtubule associated serine/threonine kinase 2 [Source:HGNC Symbol;Acc:HGNC:19035] |
| ENSG00000015479 | MATR3 | matrin 3 [Source:HGNC Symbol;Acc:HGNC:6912] |
| ENSG00000280987 | MATR3 | matrin 3 [Source:NCBI gene;Acc:9782] |
| ENSG00000153898 | MCOLN2 | mucolipin 2 [Source:HGNC Symbol;Acc:HGNC:13357] |
| ENSG00000153898 | MCOLN2 | mucolipin 2 [Source:HGNC Symbol;Acc:HGNC:13357] |
| ENSG00000153898 | MCOLN2 | mucolipin 2 [Source:HGNC Symbol;Acc:HGNC:13357] |
| ENSG00000162591 | MEGF6 | multiple EGF like domains 6 [Source:HGNC Symbol;Acc:HGNC:3232] |
| ENSG00000162591 | MEGF6 | multiple EGF like domains 6 [Source:HGNC Symbol;Acc:HGNC:3232] |
| ENSG00000162591 | MEGF6 | multiple EGF like domains 6 [Source:HGNC Symbol;Acc:HGNC:3232] |
| ENSG00000162591 | MEGF6 | multiple EGF like domains 6 [Source:HGNC Symbol;Acc:HGNC:3232] |
| None | None | None |
| None | None | None |
| None | None | None |
| ENSG00000225206 | MIR137HG | MIR137 host gene [Source:HGNC Symbol;Acc:HGNC:42871] |
| ENSG00000264406 | MIR548AP | microRNA 548ap [Source:HGNC Symbol;Acc:HGNC:43465] |
| ENSG00000269956 | MKNK1-AS1 | MKNK1 antisense RNA 1 [Source:HGNC Symbol;Acc:HGNC:44129] |
| ENSG00000142606 | MMEL1 | membrane metalloendopeptidase like 1 [Source:HGNC Symbol;Acc:HGNC:14668] |
| ENSG00000116151 | MORN1 | MORN repeat containing 1 [Source:HGNC Symbol;Acc:HGNC:25852] |
| ENSG00000116151 | MORN1 | MORN repeat containing 1 [Source:HGNC Symbol;Acc:HGNC:25852] |
| ENSG00000271723 | MROH7-TTC | MROH7-TTC4 readthrough (NMD candidate) [Source:HGNC Symbol;Acc:HGNC:49180] |
| ENSG00000271723 | MROH7-TTC | MROH7-TTC4 readthrough (NMD candidate) [Source:HGNC Symbol;Acc:HGNC:49180] |
| ENSG00000116221 | MRPL37 | mitochondrial ribosomal protein L37 [Source:HGNC Symbol;Acc:HGNC:14034] |
| ENSG00000057468 | MSH4 | mutS homolog 4 [Source:HGNC Symbol;Acc:HGNC:7327] |

| | | |
|---|---|---|
| ENSG00000057468 | MSH4 | mutS homolog 4 [Source:HGNC Symbol;Acc:HGNC:7327] |
| ENSG00000173531 | MST1 | macrophage stimulating 1 [Source:HGNC Symbol;Acc:HGNC:7380] |
| ENSG00000188786 | MTF1 | metal regulatory transcription factor 1 [Source:HGNC Symbol;Acc:HGNC:7428] |
| ENSG00000117640 | MTFR1L | mitochondrial fission regulator 1 like [Source:HGNC Symbol;Acc:HGNC:28836] |
| ENSG00000162576 | MXRA8 | matrix remodeling associated 8 [Source:HGNC Symbol;Acc:HGNC:7542] |
| ENSG00000162601 | MYSM1 | Myb like, SWIRM and MPN domains 1 [Source:HGNC Symbol;Acc:HGNC:29401] |
| ENSG00000162601 | MYSM1 | Myb like, SWIRM and MPN domains 1 [Source:HGNC Symbol;Acc:HGNC:29401] |
| ENSG00000008130 | NADK | NAD kinase [Source:HGNC Symbol;Acc:HGNC:29831] |
| ENSG00000132780 | NASP | nuclear autoantigenic sperm protein [Source:HGNC Symbol;Acc:HGNC:7644] |
| ENSG00000219481 | NBPF1 | NBPF member 1 [Source:HGNC Symbol;Acc:HGNC:26088] |
| ENSG00000219481 | NBPF1 | NBPF member 1 [Source:HGNC Symbol;Acc:HGNC:26088] |
| ENSG00000219481 | NBPF1 | NBPF member 1 [Source:HGNC Symbol;Acc:HGNC:26088] |
| ENSG00000142794 | NBPF3 | NBPF member 3 [Source:HGNC Symbol;Acc:HGNC:25076] |
| ENSG00000142794 | NBPF3 | NBPF member 3 [Source:HGNC Symbol;Acc:HGNC:25076] |
| ENSG00000270231 | NBPF8 | NBPF member 8 [Source:HGNC Symbol;Acc:HGNC:31990] |
| ENSG00000270231 | NBPF8 | NBPF member 8 [Source:HGNC Symbol;Acc:HGNC:31990] |
| ENSG00000270231 | NBPF8 | NBPF member 8 [Source:HGNC Symbol;Acc:HGNC:31990] |
| ENSG00000184454 | NCMAP | non-compact myelin associated protein [Source:HGNC Symbol;Acc:HGNC:29332] |
| ENSG00000184454 | NCMAP | non-compact myelin associated protein [Source:HGNC Symbol;Acc:HGNC:29332] |
| ENSG00000172260 | NEGR1 | neuronal growth regulator 1 [Source:HGNC Symbol;Acc:HGNC:17302] |
| ENSG00000172260 | NEGR1 | neuronal growth regulator 1 [Source:HGNC Symbol;Acc:HGNC:17302] |
| ENSG00000172260 | NEGR1 | neuronal growth regulator 1 [Source:HGNC Symbol;Acc:HGNC:17302] |
| ENSG00000172260 | NEGR1 | neuronal growth regulator 1 [Source:HGNC Symbol;Acc:HGNC:17302] |
| ENSG00000172260 | NEGR1 | neuronal growth regulator 1 [Source:HGNC Symbol;Acc:HGNC:17302] |
| ENSG00000172260 | NEGR1 | neuronal growth regulator 1 [Source:HGNC Symbol;Acc:HGNC:17302] |
| ENSG00000162599 | NFIA | nuclear factor I A [Source:HGNC Symbol;Acc:HGNC:7784] |
| ENSG00000162599 | NFIA | nuclear factor I A [Source:HGNC Symbol;Acc:HGNC:7784] |
| ENSG00000066136 | NFYC | nuclear transcription factor Y subunit gamma [Source:HGNC Symbol;Acc:HGNC:7806] |
| ENSG00000001461 | NIPAL3 | NIPA like domain containing 3 [Source:HGNC Symbol;Acc:HGNC:25233] |
| ENSG00000001461 | NIPAL3 | NIPA like domain containing 3 [Source:HGNC Symbol;Acc:HGNC:25233] |
| ENSG00000123213 | NLN | neurolysin [Source:HGNC Symbol;Acc:HGNC:16058] |
| ENSG00000188976 | NOC2L | NOC2 like nucleolar associated transcriptional repressor [Source:HGNC Symbol;Acc:HGNC:24517] |
| ENSG00000162408 | NOL9 | nucleolar protein 9 [Source:HGNC Symbol;Acc:HGNC:26265] |
| ENSG00000162408 | NOL9 | nucleolar protein 9 [Source:HGNC Symbol;Acc:HGNC:26265] |
| None | None | None |
| None | None | None |
| ENSG00000134250 | NOTCH2 | notch receptor 2 [Source:HGNC Symbol;Acc:HGNC:7882] |
| ENSG00000134250 | NOTCH2 | notch receptor 2 [Source:HGNC Symbol;Acc:HGNC:7882] |
| ENSG00000131697 | NPHP4 | nephrocystin 4 [Source:HGNC Symbol;Acc:HGNC:19104] |
| ENSG00000131697 | NPHP4 | nephrocystin 4 [Source:HGNC Symbol;Acc:HGNC:19104] |
| ENSG00000162631 | NTNG1 | netrin G1 [Source:HGNC Symbol;Acc:HGNC:23319] |
| ENSG00000118733 | OLFM3 | olfactomedin 3 [Source:HGNC Symbol;Acc:HGNC:17990] |
| ENSG00000142623 | PADI1 | peptidyl arginine deiminase 1 [Source:HGNC Symbol;Acc:HGNC:18367] |
| ENSG00000117115 | PADI2 | peptidyl arginine deiminase 2 [Source:HGNC Symbol;Acc:HGNC:18341] |
| ENSG00000117115 | PADI2 | peptidyl arginine deiminase 2 [Source:HGNC Symbol;Acc:HGNC:18341] |
| ENSG00000117115 | PADI2 | peptidyl arginine deiminase 2 [Source:HGNC Symbol;Acc:HGNC:18341] |
| ENSG00000117115 | PADI2 | peptidyl arginine deiminase 2 [Source:HGNC Symbol;Acc:HGNC:18341] |
| ENSG00000142619 | PADI3 | peptidyl arginine deiminase 3 [Source:HGNC Symbol;Acc:HGNC:18337] |
| ENSG00000276747 | PADI6 | peptidyl arginine deiminase 6 [Source:HGNC Symbol;Acc:HGNC:20449] |
| ENSG00000276747 | PADI6 | peptidyl arginine deiminase 6 [Source:HGNC Symbol;Acc:HGNC:20449] |
| ENSG00000276747 | PADI6 | peptidyl arginine deiminase 6 [Source:HGNC Symbol;Acc:HGNC:20449] |
| ENSG00000009709 | PAX7 | paired box 7 [Source:HGNC Symbol;Acc:HGNC:8621] |
| ENSG00000184588 | PDE4B | phosphodiesterase 4B [Source:HGNC Symbol;Acc:HGNC:8781] |
| ENSG00000184588 | PDE4B | phosphodiesterase 4B [Source:HGNC Symbol;Acc:HGNC:8781] |
| ENSG00000179889 | PDXDC1 | pyridoxal dependent decarboxylase domain containing 1 [Source:HGNC Symbol;Acc:HGNC:28995] |
| ENSG00000049246 | PER3 | period circadian regulator 3 [Source:HGNC Symbol;Acc:HGNC:8847] |
| ENSG00000142655 | PEX14 | peroxisomal biogenesis factor 14 [Source:HGNC Symbol;Acc:HGNC:8856] |
| ENSG00000142655 | PEX14 | peroxisomal biogenesis factor 14 [Source:HGNC Symbol;Acc:HGNC:8856] |
| ENSG00000204138 | PHACTR4 | phosphatase and actin regulator 4 [Source:HGNC Symbol;Acc:HGNC:25793] |
| ENSG00000134686 | PHC2 | polyhomeotic homolog 2 [Source:HGNC Symbol;Acc:HGNC:3183] |
| ENSG00000134686 | PHC2 | polyhomeotic homolog 2 [Source:HGNC Symbol;Acc:HGNC:3183] |
| ENSG00000116793 | PHTF1 | putative homeodomain transcription factor 1 [Source:HGNC Symbol;Acc:HGNC:8939] |
| ENSG00000065243 | PKN2 | protein kinase N2 [Source:HGNC Symbol;Acc:HGNC:9406] |
| ENSG00000149527 | PLCH2 | phospholipase C eta 2 [Source:HGNC Symbol;Acc:HGNC:29037] |
| ENSG00000171680 | PLEKHG5 | pleckstrin homology and RhoGEF domain containing G5 [Source:HGNC Symbol;Acc:HGNC:29105] |
| None | None | None |
| None | None | None |
| ENSG00000040487 | SLC66A1 | solute carrier family 66 member 1 [Source:HGNC Symbol;Acc:HGNC:26001] |
| ENSG00000142611 | PRDM16 | PR/SET domain 16 [Source:HGNC Symbol;Acc:HGNC:14000] |
| ENSG00000142611 | PRDM16 | PR/SET domain 16 [Source:HGNC Symbol;Acc:HGNC:14000] |
| ENSG00000142611 | PRDM16 | PR/SET domain 16 [Source:HGNC Symbol;Acc:HGNC:14000] |
| ENSG00000116731 | PRDM2 | PR/SET domain 2 [Source:HGNC Symbol;Acc:HGNC:9347] |
| ENSG00000162409 | PRKAA2 | protein kinase AMP-activated catalytic subunit alpha 2 [Source:HGNC Symbol;Acc:HGNC:9377] |
| ENSG00000142875 | PRKACB | protein kinase cAMP-activated catalytic subunit beta [Source:HGNC Symbol;Acc:HGNC:9381] |
| ENSG00000067606 | PRKCZ | protein kinase C zeta [Source:HGNC Symbol;Acc:HGNC:9412] |
| ENSG00000169403 | PTAFR | platelet activating factor receptor [Source:HGNC Symbol;Acc:HGNC:9582] |
| ENSG00000117425 | PTCH2 | patched 2 [Source:HGNC Symbol;Acc:HGNC:9586] |
| None | None | None |

| | | |
|---|---|---|
| None | None | None |
| ENSG00000134247 | PTGFRN | prostaglandin F2 receptor inhibitor [Source:HGNC Symbol;Acc:HGNC:9601] |
| ENSG00000142949 | PTPRF | protein tyrosine phosphatase receptor type F [Source:HGNC Symbol;Acc:HGNC:9670] |
| ENSG00000142949 | PTPRF | protein tyrosine phosphatase receptor type F [Source:HGNC Symbol;Acc:HGNC:9670] |
| ENSG00000142949 | PTPRF | protein tyrosine phosphatase receptor type F [Source:HGNC Symbol;Acc:HGNC:9670] |
| ENSG00000142949 | PTPRF | protein tyrosine phosphatase receptor type F [Source:HGNC Symbol;Acc:HGNC:9670] |
| ENSG00000060656 | PTPRU | protein tyrosine phosphatase receptor type U [Source:HGNC Symbol;Acc:HGNC:9683] |
| ENSG00000169213 | RAB3B | RAB3B, member RAS oncogene family [Source:HGNC Symbol;Acc:HGNC:9778] |
| ENSG00000169213 | RAB3B | RAB3B, member RAS oncogene family [Source:HGNC Symbol;Acc:HGNC:9778] |
| ENSG00000169213 | RAB3B | RAB3B, member RAS oncogene family [Source:HGNC Symbol;Acc:HGNC:9778] |
| ENSG00000116473 | RAP1A | RAP1A, member of RAS oncogene family [Source:HGNC Symbol;Acc:HGNC:9855] |
| ENSG00000076864 | RAP1GAP | RAP1 GTPase activating protein [Source:HGNC Symbol;Acc:HGNC:9858] |
| ENSG00000076864 | RAP1GAP | RAP1 GTPase activating protein [Source:HGNC Symbol;Acc:HGNC:9858] |
| ENSG00000162521 | RBBP4 | RB binding protein 4, chromatin remodeling factor [Source:HGNC Symbol;Acc:HGNC:9887] |
| ENSG00000134597 | RBMX2 | RNA binding motif protein X-linked 2 [Source:HGNC Symbol;Acc:HGNC:24282] |
| ENSG00000134193 | REG4 | regenerating family member 4 [Source:HGNC Symbol;Acc:HGNC:22977] |
| ENSG00000142599 | RERE | arginine-glutamic acid dipeptide repeats [Source:HGNC Symbol;Acc:HGNC:9965] |
| ENSG00000142599 | RERE | arginine-glutamic acid dipeptide repeats [Source:HGNC Symbol;Acc:HGNC:9965] |
| ENSG00000142599 | RERE | arginine-glutamic acid dipeptide repeats [Source:HGNC Symbol;Acc:HGNC:9965] |
| ENSG00000142599 | RERE | arginine-glutamic acid dipeptide repeats [Source:HGNC Symbol;Acc:HGNC:9965] |
| ENSG00000142599 | RERE | arginine-glutamic acid dipeptide repeats [Source:HGNC Symbol;Acc:HGNC:9965] |
| ENSG00000142599 | RERE | arginine-glutamic acid dipeptide repeats [Source:HGNC Symbol;Acc:HGNC:9965] |
| ENSG00000142599 | RERE | arginine-glutamic acid dipeptide repeats [Source:HGNC Symbol;Acc:HGNC:9965] |
| ENSG00000188672 | RHCE | Rh blood group CcEe antigens [Source:HGNC Symbol;Acc:HGNC:10008] |
| ENSG00000117016 | RIMS3 | regulating synaptic membrane exocytosis 3 [Source:HGNC Symbol;Acc:HGNC:21292] |
| ENSG00000158286 | RNF207 | ring finger protein 207 [Source:HGNC Symbol;Acc:HGNC:32947] |
| ENSG00000187147 | RNF220 | ring finger protein 220 [Source:HGNC Symbol;Acc:HGNC:25552] |
| ENSG00000187147 | RNF220 | ring finger protein 220 [Source:HGNC Symbol;Acc:HGNC:25552] |
| ENSG00000187147 | RNF220 | ring finger protein 220 [Source:HGNC Symbol;Acc:HGNC:25552] |
| ENSG00000187147 | RNF220 | ring finger protein 220 [Source:HGNC Symbol;Acc:HGNC:25552] |
| ENSG00000185483 | ROR1 | receptor tyrosine kinase like orphan receptor 1 [Source:HGNC Symbol;Acc:HGNC:10256] |
| ENSG00000185483 | ROR1 | receptor tyrosine kinase like orphan receptor 1 [Source:HGNC Symbol;Acc:HGNC:10256] |
| ENSG00000185483 | ROR1 | receptor tyrosine kinase like orphan receptor 1 [Source:HGNC Symbol;Acc:HGNC:10256] |
| ENSG00000185483 | ROR1 | receptor tyrosine kinase like orphan receptor 1 [Source:HGNC Symbol;Acc:HGNC:10256] |
| ENSG00000117676 | RPS6KA1 | ribosomal protein S6 kinase A1 [Source:HGNC Symbol;Acc:HGNC:10430] |
| ENSG00000137996 | RTCA | RNA 3'-terminal phosphate cyclase [Source:HGNC Symbol;Acc:HGNC:17981] |
| ENSG00000020633 | RUNX3 | RUNX family transcription factor 3 [Source:HGNC Symbol;Acc:HGNC:10473] |
| ENSG00000116497 | S100PBP | S100P binding protein [Source:HGNC Symbol;Acc:HGNC:25768] |
| ENSG00000116497 | S100PBP | S100P binding protein [Source:HGNC Symbol;Acc:HGNC:25768] |
| ENSG00000187634 | SAMD11 | sterile alpha motif domain containing 11 [Source:HGNC Symbol;Acc:HGNC:28706] |
| ENSG00000156876 | SASS6 | SAS-6 centriolar assembly protein [Source:HGNC Symbol;Acc:HGNC:25403] |
| ENSG00000156876 | SASS6 | SAS-6 centriolar assembly protein [Source:HGNC Symbol;Acc:HGNC:25403] |
| ENSG00000119231 | SENP5 | SUMO specific peptidase 5 [Source:HGNC Symbol;Acc:HGNC:28407] |
| ENSG00000168528 | SERINC2 | serine incorporator 2 [Source:HGNC Symbol;Acc:HGNC:23231] |
| ENSG00000183431 | SF3A3 | splicing factor 3a subunit 3 [Source:HGNC Symbol;Acc:HGNC:10767] |
| ENSG00000118473 | SGIP1 | SH3GL interacting endocytic adaptor 1 [Source:HGNC Symbol;Acc:HGNC:25412] |
| None | None | None |
| ENSG00000162383 | SLC1A7 | solute carrier family 1 member 7 [Source:HGNC Symbol;Acc:HGNC:10945] |
| ENSG00000085491 | SLC25A24 | solute carrier family 25 member 24 [Source:HGNC Symbol;Acc:HGNC:20662] |
| ENSG00000142583 | SLC2A5 | solute carrier family 2 member 5 [Source:HGNC Symbol;Acc:HGNC:11010] |
| None | None | None |
| ENSG00000137968 | SLC44A5 | solute carrier family 44 member 5 [Source:HGNC Symbol;Acc:HGNC:28524] |
| ENSG00000137968 | SLC44A5 | solute carrier family 44 member 5 [Source:HGNC Symbol;Acc:HGNC:28524] |
| ENSG00000137968 | SLC44A5 | solute carrier family 44 member 5 [Source:HGNC Symbol;Acc:HGNC:28524] |
| ENSG00000117834 | SLC5A9 | solute carrier family 5 member 9 [Source:HGNC Symbol;Acc:HGNC:22146] |
| ENSG00000084070 | SMAP2 | small ArfGAP2 [Source:HGNC Symbol;Acc:HGNC:25082] |
| ENSG00000084070 | SMAP2 | small ArfGAP2 [Source:HGNC Symbol;Acc:HGNC:25082] |
| ENSG00000162627 | SNX7 | sorting nexin 7 [Source:HGNC Symbol;Acc:HGNC:14971] |
| ENSG00000162627 | SNX7 | sorting nexin 7 [Source:HGNC Symbol;Acc:HGNC:14971] |
| ENSG00000162627 | SNX7 | sorting nexin 7 [Source:HGNC Symbol;Acc:HGNC:14971] |
| ENSG00000162627 | SNX7 | sorting nexin 7 [Source:HGNC Symbol;Acc:HGNC:14971] |
| ENSG00000134243 | SORT1 | sortilin 1 [Source:HGNC Symbol;Acc:HGNC:11186] |
| ENSG00000134243 | SORT1 | sortilin 1 [Source:HGNC Symbol;Acc:HGNC:11186] |
| ENSG00000155761 | SPAG17 | sperm associated antigen 17 [Source:HGNC Symbol;Acc:HGNC:26620] |
| ENSG00000187144 | SPATA21 | spermatogenesis associated 21 [Source:HGNC Symbol;Acc:HGNC:28026] |
| ENSG00000132122 | SPATA6 | spermatogenesis associated 6 [Source:HGNC Symbol;Acc:HGNC:18309] |
| ENSG00000171621 | SPSB1 | splA/ryanodine receptor domain and SOCS box containing 1 [Source:HGNC Symbol;Acc:HGNC:30628] |
| None | None | None |
| ENSG00000230806 | SRGAP2-AS1 | SRGAP2 antisense RNA 1 [Source:HGNC Symbol;Acc:HGNC:40902] |
| ENSG00000230806 | SRGAP2-AS1 | SRGAP2 antisense RNA 1 [Source:HGNC Symbol;Acc:HGNC:40902] |
| ENSG00000157216 | SSBP3 | single stranded DNA binding protein 3 [Source:HGNC Symbol;Acc:HGNC:15674] |
| ENSG00000157216 | SSBP3 | single stranded DNA binding protein 3 [Source:HGNC Symbol;Acc:HGNC:15674] |
| ENSG00000117155 | SSX2IP | SSX family member 2 interacting protein [Source:HGNC Symbol;Acc:HGNC:16509] |
| ENSG00000184005 | ST6GALNAC3 | ST6 N-acetylgalactosaminide alpha-2,6-sialyltransferase 3 [Source:HGNC Symbol;Acc:HGNC:19343] |
| ENSG00000184005 | ST6GALNAC3 | ST6 N-acetylgalactosaminide alpha-2,6-sialyltransferase 3 [Source:HGNC Symbol;Acc:HGNC:19343] |
| ENSG00000184005 | ST6GALNAC3 | ST6 N-acetylgalactosaminide alpha-2,6-sialyltransferase 3 [Source:HGNC Symbol;Acc:HGNC:19343] |
| ENSG00000184005 | ST6GALNAC3 | ST6 N-acetylgalactosaminide alpha-2,6-sialyltransferase 3 [Source:HGNC Symbol;Acc:HGNC:19343] |
| ENSG00000184005 | ST6GALNAC3 | ST6 N-acetylgalactosaminide alpha-2,6-sialyltransferase 3 [Source:HGNC Symbol;Acc:HGNC:19343] |

| | | |
|---|---|---|
| ENSG00000117069 | ST6GALNAC5 | ST6 N-acetylgalactosaminide alpha-2,6-sialyltransferase 5 [Source:HGNC Symbol;Acc:HGNC:19342] |
| ENSG00000123473 | STIL | STIL centriolar assembly protein [Source:HGNC Symbol;Acc:HGNC:10879] |
| ENSG00000117632 | STMN1 | stathmin 1 [Source:HGNC Symbol;Acc:HGNC:6510] |
| ENSG00000116266 | STXBP3 | syntaxin binding protein 3 [Source:HGNC Symbol;Acc:HGNC:11446] |
| ENSG00000143028 | SYPL2 | synaptophysin like 2 [Source:HGNC Symbol;Acc:HGNC:27638] |
| ENSG00000134207 | SYT6 | synaptotagmin 6 [Source:HGNC Symbol;Acc:HGNC:18638] |
| ENSG00000134207 | SYT6 | synaptotagmin 6 [Source:HGNC Symbol;Acc:HGNC:18638] |
| ENSG00000173662 | TAS1R1 | taste 1 receptor member 1 [Source:HGNC Symbol;Acc:HGNC:14448] |
| ENSG00000179002 | TAS1R2 | taste 1 receptor member 2 [Source:HGNC Symbol;Acc:HGNC:14905] |
| ENSG00000179002 | TAS1R2 | taste 1 receptor member 2 [Source:HGNC Symbol;Acc:HGNC:14905] |
| ENSG00000092607 | TBX15 | T-box transcription factor 15 [Source:HGNC Symbol;Acc:HGNC:11594] |
| ENSG00000070759 | TESK2 | testis associated actin remodelling kinase 2 [Source:HGNC Symbol;Acc:HGNC:11732] |
| ENSG00000069702 | TGFBR3 | transforming growth factor beta receptor 3 [Source:HGNC Symbol;Acc:HGNC:11774] |
| ENSG00000069702 | TGFBR3 | transforming growth factor beta receptor 3 [Source:HGNC Symbol;Acc:HGNC:11774] |
| ENSG00000069702 | TGFBR3 | transforming growth factor beta receptor 3 [Source:HGNC Symbol;Acc:HGNC:11774] |
| ENSG00000054118 | THRAP3 | thyroid hormone receptor associated protein 3 [Source:HGNC Symbol;Acc:HGNC:22964] |
| ENSG00000162542 | TMCO4 | transmembrane and coiled-coil domains 4 [Source:HGNC Symbol;Acc:HGNC:27393] |
| ENSG00000162542 | TMCO4 | transmembrane and coiled-coil domains 4 [Source:HGNC Symbol;Acc:HGNC:27393] |
| ENSG00000162542 | TMCO4 | transmembrane and coiled-coil domains 4 [Source:HGNC Symbol;Acc:HGNC:27393] |
| ENSG00000171729 | TMEM51 | transmembrane protein 51 [Source:HGNC Symbol;Acc:HGNC:25488] |
| ENSG00000171729 | TMEM51 | transmembrane protein 51 [Source:HGNC Symbol;Acc:HGNC:25488] |
| ENSG00000171729 | TMEM51 | transmembrane protein 51 [Source:HGNC Symbol;Acc:HGNC:25488] |
| ENSG00000175147 | TMEM51-AS | TMEM51 antisense RNA 1 [Source:HGNC Symbol;Acc:HGNC:26301] |
| None | None | None |
| None | None | None |
| ENSG00000116209 | TMEM59 | transmembrane protein 59 [Source:HGNC Symbol;Acc:HGNC:1239] |
| ENSG00000120949 | TNFRSF8 | TNF receptor superfamily member 8 [Source:HGNC Symbol;Acc:HGNC:11923] |
| ENSG00000078900 | TP73 | tumor protein p73 [Source:HGNC Symbol;Acc:HGNC:12003] |
| ENSG00000269113 | TRABD2B | TraB domain containing 2B [Source:HGNC Symbol;Acc:HGNC:44200] |
| ENSG00000269113 | TRABD2B | TraB domain containing 2B [Source:HGNC Symbol;Acc:HGNC:44200] |
| ENSG00000197323 | TRIM33 | tripartite motif containing 33 [Source:HGNC Symbol;Acc:HGNC:16290] |
| ENSG00000197323 | TRIM33 | tripartite motif containing 33 [Source:HGNC Symbol;Acc:HGNC:16290] |
| ENSG00000116525 | TRIM62 | tripartite motif containing 62 [Source:HGNC Symbol;Acc:HGNC:25574] |
| ENSG00000043514 | TRIT1 | tRNA isopentenyltransferase 1 [Source:HGNC Symbol;Acc:HGNC:20286] |
| ENSG00000134198 | TSPAN2 | tetraspanin 2 [Source:HGNC Symbol;Acc:HGNC:20659] |
| ENSG00000006555 | TTC22 | tetratricopeptide repeat domain 22 [Source:HGNC Symbol;Acc:HGNC:26067] |
| ENSG00000085831 | TTC39A | tetratricopeptide repeat domain 39A [Source:HGNC Symbol;Acc:HGNC:18657] |
| ENSG00000159247 | TUBBP5 | tubulin beta pseudogene 5 [Source:HGNC Symbol;Acc:HGNC:23674] |
| ENSG00000159247 | TUBBP5 | tubulin beta pseudogene 5 [Source:HGNC Symbol;Acc:HGNC:23674] |
| ENSG00000158062 | UBXN11 | UBX domain protein 11 [Source:HGNC Symbol;Acc:HGNC:30600] |
| ENSG00000158062 | UBXN11 | UBX domain protein 11 [Source:HGNC Symbol;Acc:HGNC:30600] |
| ENSG00000162402 | USP24 | ubiquitin specific peptidase 24 [Source:HGNC Symbol;Acc:HGNC:12623] |
| ENSG00000162402 | USP24 | ubiquitin specific peptidase 24 [Source:HGNC Symbol;Acc:HGNC:12623] |
| ENSG00000090686 | USP48 | ubiquitin specific peptidase 48 [Source:HGNC Symbol;Acc:HGNC:18533] |
| ENSG00000134215 | VAV3 | vav guanine nucleotide exchange factor 3 [Source:HGNC Symbol;Acc:HGNC:12659] |
| ENSG00000134215 | VAV3 | vav guanine nucleotide exchange factor 3 [Source:HGNC Symbol;Acc:HGNC:12659] |
| ENSG00000134215 | VAV3 | vav guanine nucleotide exchange factor 3 [Source:HGNC Symbol;Acc:HGNC:12659] |
| ENSG00000134215 | VAV3 | vav guanine nucleotide exchange factor 3 [Source:HGNC Symbol;Acc:HGNC:12659] |
| ENSG00000230489 | VAV3-AS1 | VAV3 antisense RNA 1 [Source:HGNC Symbol;Acc:HGNC:40608] |
| ENSG00000048707 | VPS13D | vacuolar protein sorting 13 homolog D [Source:HGNC Symbol;Acc:HGNC:23595] |
| ENSG00000134258 | VTCN1 | V-set domain containing T cell activation inhibitor 1 [Source:HGNC Symbol;Acc:HGNC:28873] |
| ENSG00000134258 | VTCN1 | V-set domain containing T cell activation inhibitor 1 [Source:HGNC Symbol;Acc:HGNC:28873] |
| ENSG00000116874 | WARS2 | tryptophanyl tRNA synthetase 2, mitochondrial [Source:HGNC Symbol;Acc:HGNC:12730] |
| ENSG00000158195 | WASF2 | WASP family member 2 [Source:HGNC Symbol;Acc:HGNC:12733] |
| ENSG00000158195 | WASF2 | WASP family member 2 [Source:HGNC Symbol;Acc:HGNC:12733] |
| ENSG00000185596 | WASH3P | WASP family homolog 3, pseudogene [Source:HGNC Symbol;Acc:HGNC:24362] |
| ENSG00000162643 | WDR63 | WD repeat domain 63 [Source:HGNC Symbol;Acc:HGNC:30711] |
| ENSG00000162643 | WDR63 | WD repeat domain 63 [Source:HGNC Symbol;Acc:HGNC:30711] |
| None | None | None |
| ENSG00000152763 | WDR78 | WD repeat domain 78 [Source:HGNC Symbol;Acc:HGNC:26252] |
| ENSG00000116809 | ZBTB17 | zinc finger and BTB domain containing 17 [Source:HGNC Symbol;Acc:HGNC:12936] |
| ENSG00000157077 | ZFYVE9 | zinc finger FYVE-type containing 9 [Source:HGNC Symbol;Acc:HGNC:6775] |
| ENSG00000157077 | ZFYVE9 | zinc finger FYVE-type containing 9 [Source:HGNC Symbol;Acc:HGNC:6775] |
| ENSG00000157077 | ZFYVE9 | zinc finger FYVE-type containing 9 [Source:HGNC Symbol;Acc:HGNC:6775] |
| ENSG00000066185 | ZMYND12 | zinc finger MYND-type containing 12 [Source:HGNC Symbol;Acc:HGNC:21192] |
| ENSG00000162664 | ZNF326 | zinc finger protein 326 [Source:HGNC Symbol;Acc:HGNC:14104] |
| ENSG00000172748 | ZNF596 | zinc finger protein 596 [Source:HGNC Symbol;Acc:HGNC:27268] |
| ENSG00000162415 | ZSWIM5 | zinc finger SWIM-type containing 5 [Source:HGNC Symbol;Acc:HGNC:29299] |
| ENSG00000162378 | ZYG11B | zyg-11 family member B, cell cycle regulator [Source:HGNC Symbol;Acc:HGNC:25820] |
| ENSG00000036549 | AC118549.1 | zinc finger ZZ-type containing 3 [Source:NCBI gene;Acc:26009] |
| ENSG00000036549 | AC118549.1 | zinc finger ZZ-type containing 3 [Source:NCBI gene;Acc:26009] |
