## Supplementary material for "Genetic Mutations Associated with Hormone-Positive Breast Cancer in Ethiopian Women": Table S2

**Results from the GSEA enrichments**

| Gene Set Name | # Genes in Gene Set (K) | Description | # Genes in Overlap (k) | k/K | p-value | FDR q-value |
|---|---|---|---|---|---|---|
| GO_HYDROLASE_ACTIVITY_ACTING_ON_CARBON_NITROGEN_BUT_NOT_PEPTIDE_BONDS_IN_LINEAR_AMIDINES | 11 | Catalysis of the hydrolysis of any non-peptide carbon-nitrogen bond in a linear amidine, a compound of the form R-C(=NH)-NH2. [ISBN:0198506732] | 3 | 0.3 | 2.96E-06 | 3.06E-02 |
| REACTOME_GLUCAGON_LIKE_PEPTIDE_1_GLP1_REGULATES_INSULIN_SECRETION | 42 | Glucagon-like Peptide-1 (GLP1) regulates insulin secretion | 4 | 0.1 | 4.86E-06 | 3.06E-02 |
| VEGF_A_UP.V1_DN | 192 | Genes down-regulated in HUVEC cells (endothelium) by treatment with VEGFA [Gene ID=7422]. | 6 | 0 | 1.31E-05 | 3.27E-02 |
| GO_CENTRAL_NERVOUS_SYSTEM_DEVELOPMENT | 995 | The process whose specific outcome is the progression of the central nervous system over time, from its formation to the mature structure. The central nervous system is the core nervous system that serves an integrating and coordinating function. In vertebrates it consists of the brain and spinal cord. In those invertebrates with a central nervous system it typically consists of a brain, cerebral ganglia and a nerve cord. [GOC:bf, GOC:jid, ISBN:0582227089] | 12 | 0 | 1.36E-05 | 3.27E-02 |
| GO_PLASMA_MEMBRANE_PROTEIN_COMPLEX | 680 | Any protein complex that is part of the plasma membrane. [GOC:dos] | 10 | 0 | 1.40E-05 | 3.27E-02 |
| GO_RESPONSE_TO_INTERFERON_GAMMA | 198 | Any process that results in a change in state or activity of a cell or an organism (in terms of movement, secretion, enzyme production, gene expression, etc.) as a result of an interferon-gamma stimulus. Interferon-gamma is also known as type II interferon. [GOC:add, ISBN:0126896631, PMID:15546383] | 6 | 0 | 1.55E-05 | 3.27E-02 |
| GO_SMALL_GTPASE_MEDIATED_SIGNAL_TRANSDUCTION | 569 | Any series of molecular signals in which a small monomeric GTPase relays one or more of the signals. [GOC:mah] | 9 | 0 | 2.18E-05 | 3.68E-02 |
| REACTOME_PRESYNAPTIC_FUNCTION_OF_KAINATE_RECEPTORS | 21 | Presynaptic function of Kainate receptors | 3 | 0.1 | 2.34E-05 | 3.68E-02 |
| GO_RAS_PROTEIN_SIGNAL_TRANSDUCTION | 447 | A series of molecular signals within the cell that are mediated by a member of the Ras superfamily of proteins switching to a GTP-bound active state. [GOC:bf] | 8 | 0 | 2.67E-05 | 3.74E-02 |
| GO_HEAD_DEVELOPMENT | 774 | The biological process whose specific outcome is the progression of a head from an initial condition to its mature state. The head is the anterior-most division of the body. [GOC:dph] | 10 | 0 | 4.19E-05 | 4.91E-02 |
