## Supplementary material for "Genetic Mutations Associated with Hormone-Positive Breast Cancer in Ethiopian Women": Table S3

| Ensembl_ID | geneName | description |
|---|---|---|
| ENSG00000118733 | OLFM3 | olfactomedin 3 [Source:HGNC Symbol;Acc:HGNC:17990] |
| ENSG00000116874 | WARS2 | tryptophanyl tRNA synthetase 2, mitochondrial [Source:HGNC Symbol;Acc:HGNC:12730] |
| ENSG00000172260 | NEGR1 | neuronal growth regulator 1 [Source:HGNC Symbol;Acc:HGNC:17302] |
| ENSG00000162598 | C1orf87 | chromosome 1 open reading frame 87 [Source:HGNC Symbol;Acc:HGNC:28547] |
| ENSG00000171680 | PLEKHG5 | pleckstrin homology and RhoGEF domain containing G5 [Source:HGNC Symbol;Acc:HGNC:29105] |
| ENSG00000116473 | RAP1A | RAP1A, member of RAS oncogene family [Source:HGNC Symbol;Acc:HGNC:9855] |
| ENSG00000162378 | ZYG11B | zyg-11 family member B, cell cycle regulator [Source:HGNC Symbol;Acc:HGNC:25820] |
| ENSG00000143079 | CTTNBP2NL | CTTNBP2 N-terminal like [Source:HGNC Symbol;Acc:HGNC:25330] |
| ENSG00000153904 | DDAH1 | dimethylarginine dimethylaminohydrolase 1 [Source:HGNC Symbol;Acc:HGNC:2715] |
| ENSG00000117643 | MAN1C1 | mannosidase alpha class 1C member 1 [Source:HGNC Symbol;Acc:HGNC:19080] |
| ENSG00000116497 | S100PBP | S100P binding protein [Source:HGNC Symbol;Acc:HGNC:25768] |
| ENSG00000197429 | IPP | intracisternal A particle-promoted polypeptide [Source:HGNC Symbol;Acc:HGNC:6108] |
| ENSG00000016490 | CLCA1 | chloride channel accessory 1 [Source:HGNC Symbol;Acc:HGNC:2015] |
| ENSG00000134193 | REG4 | regenerating family member 4 [Source:HGNC Symbol;Acc:HGNC:22977] |
| ENSG00000142875 | PRKACB | protein kinase cAMP-activated catalytic subunit beta [Source:HGNC Symbol;Acc:HGNC:9381] |
| ENSG00000123472 | ATPAF1 | ATP synthase mitochondrial F1 complex assembly factor 1 [Source:HGNC Symbol;Acc:HGNC:18803] |
| ENSG00000185104 | FAF1 | Fas associated factor 1 [Source:HGNC Symbol;Acc:HGNC:3578] |
| ENSG00000117115 | PADI2 | peptidyl arginine deiminase 2 [Source:HGNC Symbol;Acc:HGNC:18341] |
| ENSG00000219073 | CELA3B | chymotrypsin like elastase family member 3B [Source:HGNC Symbol;Acc:HGNC:15945] |
| ENSG00000097021 | ACOT7 | acyl-CoA thioesterase 7 [Source:HGNC Symbol;Acc:HGNC:24157] |
| ENSG00000203965 | EFCAB7 | EF-hand calcium binding domain 7 [Source:HGNC Symbol;Acc:HGNC:29379] |
| ENSG00000276747 | PADI6 | peptidyl arginine deiminase 6 [Source:HGNC Symbol;Acc:HGNC:20449] |
| ENSG00000043514 | TRIT1 | tRNA isopentenyltransferase 1 [Source:HGNC Symbol;Acc:HGNC:20286] |
| ENSG00000162434 | JAK1 | Janus kinase 1 [Source:HGNC Symbol;Acc:HGNC:6190] |
| ENSG00000162594 | IL23R | interleukin 23 receptor [Source:HGNC Symbol;Acc:HGNC:19100] |
| ENSG00000136720 | HS6ST1 | heparan sulfate 6-O-sulfotransferase 1 [Source:HGNC Symbol;Acc:HGNC:5201] |
| ENSG00000118473 | SGIP1 | SH3 domain GRB2 like endophilin interacting protein 1 [Source:HGNC Symbol;Acc:HGNC:25412] |
| ENSG00000157211 | CDCP2 | CUB domain containing protein 2 [Source:HGNC Symbol;Acc:HGNC:27297] |
| ENSG00000162664 | ZNF326 | zinc finger protein 326 [Source:HGNC Symbol;Acc:HGNC:14104] |
| ENSG00000186094 | AGBL4 | ATP/GTP binding protein like 4 [Source:HGNC Symbol;Acc:HGNC:25892] |
| ENSG00000162482 | AKR7A3 | aldo-keto reductase family 7 member A3 [Source:HGNC Symbol;Acc:HGNC:390] |
| ENSG00000174574 | AKIRIN1 | akirin 1 [Source:HGNC Symbol;Acc:HGNC:25744] |
| ENSG00000134597 | RBMX2 | RNA binding motif protein X-linked 2 [Source:HGNC Symbol;Acc:HGNC:24282] |
| ENSG00000078900 | TP73 | tumor protein p73 [Source:HGNC Symbol;Acc:HGNC:12003] |
| ENSG00000185436 | IFNLR1 | interferon lambda receptor 1 [Source:HGNC Symbol;Acc:HGNC:18584] |
| ENSG00000116209 | TMEM59 | transmembrane protein 59 [Source:HGNC Symbol;Acc:HGNC:1239] |
| ENSG00000184454 | NCMAP | non-compact myelin associated protein [Source:HGNC Symbol;Acc:HGNC:29332] |
| ENSG00000183347 | GBP6 | guanylate binding protein family member 6 [Source:HGNC Symbol;Acc:HGNC:25395] |
| ENSG00000215704 | CELA2B | chymotrypsin like elastase family member 2B [Source:HGNC Symbol;Acc:HGNC:29995] |
| ENSG00000188976 | NOC2L | NOC2 like nucleolar associated transcriptional repressor [Source:HGNC Symbol;Acc:HGNC:24517] |
| ENSG00000204138 | PHACTR4 | phosphatase and actin regulator 4 [Source:HGNC Symbol;Acc:HGNC:25793] |
| ENSG00000116525 | TRIM62 | tripartite motif containing 62 [Source:HGNC Symbol;Acc:HGNC:25574] |
| ENSG00000117640 | MTFR1L | mitochondrial fission regulator 1 like [Source:HGNC Symbol;Acc:HGNC:28836] |
| ENSG00000142655 | PEX14 | peroxisomal biogenesis factor 14 [Source:HGNC Symbol;Acc:HGNC:8856] |
| ENSG00000036549 | None | None |
| ENSG00000197323 | TRIM33 | tripartite motif containing 33 [Source:HGNC Symbol;Acc:HGNC:16290] |
| ENSG00000122483 | CCDC18 | coiled-coil domain containing 18 [Source:HGNC Symbol;Acc:HGNC:30370] |
| ENSG00000169598 | DFFB | DNA fragmentation factor subunit beta [Source:HGNC Symbol;Acc:HGNC:2773] |
| ENSG00000004487 | KDM1A | lysine demethylase 1A [Source:HGNC Symbol;Acc:HGNC:29079] |
| ENSG00000125703 | ATG4C | autophagy related 4C cysteine peptidase [Source:HGNC Symbol;Acc:HGNC:16040] |
| ENSG00000088280 | ASAP3 | ArfGAP with SH3 domain, ankyrin repeat and PH domain 3 [Source:HGNC Symbol;Acc:HGNC:14987] |
| ENSG00000157077 | ZFYVE9 | zinc finger FYVE-type containing 9 [Source:HGNC Symbol;Acc:HGNC:6775] |
| ENSG00000090686 | USP48 | ubiquitin specific peptidase 48 [Source:HGNC Symbol;Acc:HGNC:18533] |
| ENSG00000142583 | SLC2A5 | solute carrier family 2 member 5 [Source:HGNC Symbol;Acc:HGNC:11010] |
| ENSG00000142794 | NBPF3 | NBPF member 3 [Source:HGNC Symbol;Acc:HGNC:25076] |
| ENSG00000162390 | ACOT11 | acyl-CoA thioesterase 11 [Source:HGNC Symbol;Acc:HGNC:18156] |
| ENSG00000188641 | DPYD | dihydropyrimidine dehydrogenase [Source:HGNC Symbol;Acc:HGNC:3012] |
| ENSG00000086015 | MAST2 | microtubule associated serine/threonine kinase 2 [Source:HGNC Symbol;Acc:HGNC:19035] |
| ENSG00000137960 | GIPC2 | GIPC PDZ domain containing family member 2 [Source:HGNC Symbol;Acc:HGNC:18177] |
| ENSG00000020633 | RUNX3 | runt related transcription factor 3 [Source:HGNC Symbol;Acc:HGNC:10473] |

| | | |
|---|---|---|
| ENSG00000132854 | KANK4 | KN motif and ankyrin repeat domains 4 [Source:HGNC Symbol;Acc:HGNC:27263] |
| ENSG00000137996 | RTCA | RNA 3'-terminal phosphate cyclase [Source:HGNC Symbol;Acc:HGNC:17981] |
| ENSG00000153898 | MCOLN2 | mucolipin 2 [Source:HGNC Symbol;Acc:HGNC:13357] |
| ENSG00000085831 | TTC39A | tetratricopeptide repeat domain 39A [Source:HGNC Symbol;Acc:HGNC:18657] |
| ENSG00000187634 | SAMD11 | sterile alpha motif domain containing 11 [Source:HGNC Symbol;Acc:HGNC:28706] |
| ENSG00000070785 | EIF2B3 | eukaryotic translation initiation factor 2B subunit gamma [Source:HGNC Symbol;Acc:HGNC:3259] |
| ENSG00000040487 | PQLC2 | PQ loop repeat containing 2 [Source:HGNC Symbol;Acc:HGNC:26001] |
| ENSG00000171729 | TMEM51 | transmembrane protein 51 [Source:HGNC Symbol;Acc:HGNC:25488] |
| ENSG00000116809 | ZBTB17 | zinc finger and BTB domain containing 17 [Source:HGNC Symbol;Acc:HGNC:12936] |
| ENSG00000119231 | SENP5 | SUMO specific peptidase 5 [Source:HGNC Symbol;Acc:HGNC:28407] |
| ENSG00000116641 | DOCK7 | dedicator of cytokinesis 7 [Source:HGNC Symbol;Acc:HGNC:19190] |
| ENSG00000197312 | DDI2 | DNA damage inducible 1 homolog 2 [Source:HGNC Symbol;Acc:HGNC:24578] |
| ENSG00000066136 | NFYC | nuclear transcription factor Y subunit gamma [Source:HGNC Symbol;Acc:HGNC:7806] |
| ENSG00000172456 | FGGY | FGGY carbohydrate kinase domain containing [Source:HGNC Symbol;Acc:HGNC:25610] |
| ENSG00000078369 | GNB1 | G protein subunit beta 1 [Source:HGNC Symbol;Acc:HGNC:4396] |
| ENSG00000085491 | SLC25A24 | solute carrier family 25 member 24 [Source:HGNC Symbol;Acc:HGNC:20662] |
| ENSG00000009709 | PAX7 | paired box 7 [Source:HGNC Symbol;Acc:HGNC:8621] |
| ENSG00000158195 | WASF2 | WAS protein family member 2 [Source:HGNC Symbol;Acc:HGNC:12733] |
| ENSG00000116793 | PHTF1 | putative homeodomain transcription factor 1 [Source:HGNC Symbol;Acc:HGNC:8939] |
| ENSG00000154027 | AK5 | adenylate kinase 5 [Source:HGNC Symbol;Acc:HGNC:365] |
| ENSG00000162433 | AK4 | adenylate kinase 4 [Source:HGNC Symbol;Acc:HGNC:363] |
| ENSG00000130762 | ARHGEF16 | Rho guanine nucleotide exchange factor 16 [Source:HGNC Symbol;Acc:HGNC:15515] |
| ENSG00000132122 | SPATA6 | spermatogenesis associated 6 [Source:HGNC Symbol;Acc:HGNC:18309] |
| ENSG00000169213 | RAB3B | RAB3B, member RAS oncogene family [Source:HGNC Symbol;Acc:HGNC:9778] |
| ENSG00000117419 | ERI3 | ERI1 exoribonuclease family member 3 [Source:HGNC Symbol;Acc:HGNC:17276] |
| ENSG00000143028 | SYPL2 | synaptophysin like 2 [Source:HGNC Symbol;Acc:HGNC:27638] |
| ENSG00000123473 | STIL | STIL, centriolar assembly protein [Source:HGNC Symbol;Acc:HGNC:10879] |
| ENSG00000116151 | MORN1 | MORN repeat containing 1 [Source:HGNC Symbol;Acc:HGNC:25852] |
| ENSG00000172748 | ZNF596 | zinc finger protein 596 [Source:HGNC Symbol;Acc:HGNC:27268] |
| ENSG00000162494 | LRRC38 | leucine rich repeat containing 38 [Source:HGNC Symbol;Acc:HGNC:27005] |
| ENSG00000169403 | PTAFR | platelet activating factor receptor [Source:HGNC Symbol;Acc:HGNC:9582] |
| ENSG00000174332 | GLIS1 | GLIS family zinc finger 1 [Source:HGNC Symbol;Acc:HGNC:29525] |
| ENSG00000159592 | GPBP1L1 | GC-rich promoter binding protein 1 like 1 [Source:HGNC Symbol;Acc:HGNC:28843] |
| ENSG00000001461 | NIPAL3 | NIPA like domain containing 3 [Source:HGNC Symbol;Acc:HGNC:25233] |
| ENSG00000172380 | GNG12 | G protein subunit gamma 12 [Source:HGNC Symbol;Acc:HGNC:19663] |
| ENSG00000049246 | PER3 | period circadian regulator 3 [Source:HGNC Symbol;Acc:HGNC:8847] |
| ENSG00000158062 | UBXN11 | UBX domain protein 11 [Source:HGNC Symbol;Acc:HGNC:30600] |
| ENSG00000116266 | STXBP3 | syntaxin binding protein 3 [Source:HGNC Symbol;Acc:HGNC:11446] |
| ENSG00000117155 | SSX2IP | SSX family member 2 interacting protein [Source:HGNC Symbol;Acc:HGNC:16509] |
| ENSG00000162631 | NTNG1 | netrin G1 [Source:HGNC Symbol;Acc:HGNC:23319] |
| ENSG00000163873 | GRIK3 | glutamate ionotropic receptor kainate type subunit 3 [Source:HGNC Symbol;Acc:HGNC:4581] |
| ENSG00000117054 | ACADM | acyl-CoA dehydrogenase medium chain [Source:HGNC Symbol;Acc:HGNC:89] |
| ENSG00000157193 | LRP8 | LDL receptor related protein 8 [Source:HGNC Symbol;Acc:HGNC:6700] |
